# Cost and Cost-Effectiveness of Digital Technologies for Support of Tuberculosis Treatment Adherence: A Systematic Review

**DOI:** 10.1101/2024.05.24.24307907

**Authors:** Cedric Kafie, Mona Salaheldin Mohamed, Miranda Zary, Chimweta Ian Chilala, Shruti Bahukudumbi, Genevieve Gore, Nicola Foster, Katherine Fielding, Ramnath Subbaraman, Kevin Schwartzman

## Abstract

**Background:** Digital adherence technologies (DATs) may provide a patient-centered approach for supporting tuberculosis (TB) medication adherence and improving treatment outcomes. We synthesized evidence addressing costs and cost-effectiveness of DATs to support TB treatment.

**Methods:** A systematic review (PROSPERO-CRD42022313531) identified relevant literature from January 2000-April 2023 in MEDLINE, Embase, CENTRAL, CINAHL, Web of Science along with preprints from medRxiv, Europe PMC and clinicaltrials.gov. Studies with observational, experimental, or quasi-experimental designs (minimum 20 participants) and modelling studies reporting quantitative data on the cost or cost-effectiveness of DATs for TB infection or disease treatment were included. Study characteristics, cost and cost-effectiveness outcomes were extracted.

**Results:** Of 3,619 titles identified by our systematic search, 29 studies met inclusion criteria, of which 9 addressed cost-effectiveness. DATs included SMS reminders, phone-based technologies, digital pillboxes, ingestible sensors, and video observed treatment (VOT). VOT was the most extensively studied (16 studies) and was generally cost saving when compared to healthcare provider directly observed therapy (DOT), particularly when costs to patients were included--though findings were largely from high-income countries. Cost-effectiveness findings were highly variable, ranging from no clinical effect in one study (SMS), to greater effectiveness with concurrent cost savings (VOT) in others. Only 8 studies adequately reported at least 80% of the elements required by CHEERS, a standard reporting checklist for health economic evaluations.

**Conclusion:** DATs may be cost-saving or cost-effective compared to healthcare provider DOT, particularly in high-income settings. However, more data of higher quality are needed, notably in lower- and middle-income countries which have the greatest TB burden.

**KEY MESSAGES:** *What is already known on this topic:* Digital adherence technologies (DATs) can provide a less intrusive, and potentially less resource-intensive way to monitor and support tuberculosis treatment adherence, as compared to traditional direct observation. To date, there is limited information about the cost and cost-effectiveness of these technologies in diverse care settings.

*What this study adds:* Our comprehensive review of available studies shows that some DATs like video-observed therapy can be cost-saving, particularly in higher-income countries, and especially when patient costs are considered.

*How this study might affect research, practice or policy:* While program savings related to some DATS will likely offset their initial costs in higher-income settings, more evidence is needed from lower-income settings where the TB burden is highest. Costing studies should also more rigorously account for all relevant costs, including those to patients.

## BACKGROUND

One-quarter of the world’s population is believed to have been infected with *Mycobacterium tuberculosis* [1]. In 2022, an estimated 10.6 million people fell ill with tuberculosis (TB) disease worldwide and a total of 1.3 million people died from TB [2].

Treatment for TB disease typically involves multiple antibiotics for at least six months, with a four-month regimen recently introduced in some settings [3]. Poor adherence to antituberculosis treatment may lead to treatment failure and relapse. Directly observed therapy (DOT), where an observer witnesses all or most doses, is commonly instituted with the goal of improving adherence but is seen as both coercive and resource-intensive [4]. Particularly when administered by healthcare providers, DOT often requires frequent travel, time off work, and additional childcare expenses, that can place a large financial and emotional burden on persons with TB.

Tuberculosis infection (TBI), the state of infection with *Mycobacterium tuberculosis* without any clinical symptoms, radiographic progression, or detectable bacteria, is usually treated for three to nine months with one or more antituberculosis antibiotics to clear the infection and prevent the development of active disease. Suboptimal adherence substantially limits the individual and population health benefits of such preventive treatment.

Digital adherence technologies (DATs) may facilitate more patient-centric approaches for monitoring TB medication adherence than existing directly observed therapy (DOT) models [5]. DAT interventions include smartphone-based technologies, SMS or video-supported treatment, digital pillboxes, and ingestible sensors that aim to monitor, and improve adherence to TB treatment.

Technologies that reduce travel and time requirements could produce significant cost savings while improving the treatment experience for persons with TB. However, these devices can carry significant technology costs, which are a particular challenge in lower-income settings. In addition, their impact on clinical outcomes varies by technology, intervention approach, and setting. Depending on the setting and care model, some reports have suggested that DAT-based treatment may be cost-effective or even cost-saving relative to the standard of care (which often varies), but these findings have been inconsistent [6].

We conducted a systematic review, to summarize existing evidence addressing the cost and cost-effectiveness of DATs for TB disease and TB infection.

## METHODS

The protocol for this systematic review was registered in PROSPERO, the International Prospective Register of Systematic Reviews (CRD42022313531) [7] and is summarized below.

### Search and screening strategy

The search was conducted on April 25, 2023 (updated from April 14, 2022) and included reports published in MEDLINE/Ovid, Embase/Ovid, CENTRAL/Wiley, CINAHL, and Web of Science Core Collection, from January 1, 2000 – April 14, 2023. We also searched Europe PMC for pre-prints (including medRxiv) and clinicaltrials.gov for unpublished clinical trials and investigators of interest. Key search terms related to TB (active or latent), digital technologies (such as mobile phone, smartphone, video observation, medication monitors, and text messaging), and cost (such as cost, economic cost-effectiveness, incremental cost-effectiveness ratio [ICER]). A complete list of search terms can be found in supplemental section S1. The database search was conducted by a health librarian (GG). Separately, a hand search was conducted through the Union World Conference on Lung Health conference proceedings for relevant abstracts on DATs and costs from 2004 to 2022 inclusively. There were no language restrictions.

### Inclusion/exclusion criteria

Studies were included if they reported quantitative cost, budget impact, or cost-effectiveness estimates for the use of DATs for TB treatment support (e.g. pure cost descriptions, incremental cost comparisons, cost per health gain, etc). The minimum number of participants required to use the DAT was 20 except for modelling studies, which typically reflected hypothetical cohorts and cost inputs from a variety of sources. DAT interventions included but were not limited to smartphone-based technologies, SMS or video-supported treatment, digital pillboxes, and ingestible sensors. Studies had to involve DAT use to support treatment adherence in individuals diagnosed and treated for TB disease or infection, including persons generally at higher risk of unfavorable outcomes (e.g., those with drug-resistant TB, persons living with HIV). A full case definition for DATs is included in supplemental section S2.

Eligible study designs included randomized controlled trials, quasi-experimental trials, observational studies, and modelling studies. Articles were excluded if the technology was not used to improve TB treatment adherence; we also excluded review articles, editorials, commentaries, news articles, and protocols, as well as abstracts other than those from the Union World Conference on Lung Health. Relevant grey literature publications (such as preprints, ministry reports, technical papers) were eligible if they met all inclusion criteria.

### Study selection

After de-duplication using EndNote [8], each title and abstract was screened independently (blinded) by at least two reviewers (among CK, MS, MZ, CC, and SB) using Rayyan.ai [9] to establish whether the publication in question potentially addressed DATs for TB treatment support and potentially met the other inclusion criteria. Studies for possible inclusion then underwent independent full-text review by two reviewers, for eligibility according to the detailed criteria above. Conflicts during each stage were resolved by consensus, with inclusion of a third senior reviewer when needed (KF, RS, and KS). All publications cited by the included articles, and all publications that cited them (using Google Scholar [10]) were also screened for inclusion.

### Data extraction

After identifying all eligible studies, their data were extracted into a standard template in Excel [11] by two independent reviewers (among CK, MS, and MZ) and subsequently compared. Any conflicts were resolved by consensus, and by discussion with a third reviewer when necessary. Extracted data included detailed information on the study characteristics, study design, study setting (inpatient or outpatient), participant characteristics, DAT used, intervention duration, standard of care (comparator), and any important gaps noted by the reviewers. The total costs (for the DAT and any comparator) and cost effectiveness estimates provided by the reports were extracted, as was an inventory of the cost components included in the total (e.g. equipment, personnel time, etc). Supplemental Table A1, lists and describes the types of costs tabulated for each study, whenever available. Studies that reported multiple sensitivity analyses and scale-up scenarios were noted but only the base case results were extracted.

We also extracted all available information on the scope, frequency (e.g. daily) and mode (e.g. self-administered therapy [SAT] or DOT) of the comparator. Whenever DOT was used as a comparator for a digital technology, we also noted the method of delivery (e.g. clinic or field-based). While specifics varied, we noted three main models for DOT delivery. Clinic DOT required the person taking TB treatment to attend a central facility such as a clinic, a hospital, or a prison for observation. Field DOT required health workers to travel to the person’s home, workplace, or other community location for dosing observation. Family DOT allowed observation by designated treatment supporters among family members or close friends. We noted studies where the mode for DOT was not further characterized as “DOT—not reported” i.e. DOT-NR.

### Data synthesis

In our primary analysis, we expressed costs in “international” dollars based on purchasing power parity (PPP), reflecting equivalent purchasing power in each study setting to that provided by a US dollar in the US. [12]^1^. This enhances comparability across diverse settings, and attenuates distortion from abrupt fluctuations in market exchange rates (e.g. if a currency is revalued) [13]. It is particularly relevant for local decisionmakers and funders, as opposed to international donors. Strictly speaking, the use of PPP may be best suited for non-tradable goods, e.g. local labor. However, in DAT cost studies, labor and domestic transport are often the major component, and a detailed breakdown between tradable and non-tradable goods may not be available.

Hence costs from countries other than the US were first inflated to 2022 costs using the local GDP implicit price deflator as recommended for non-traded goods [13,14] and then converted to international dollars using PPP estimates reported by the International Monetary Fund (IMF) [15]. US-based costs were inflated to 2022 dollars using the US GDP deflator from the same IMF dataset [15].

We also performed a sensitivity analysis where all costs were converted to US dollars using average annual market exchange rates from the IMF [16]. When not otherwise reported, currency year was assumed to be the publication year, and the average market exchange rates for that year were used for currency conversion. A list of all factors used for cost adjustment is provided supplemental Table A22.

We grouped cost results by DAT and tabulated the costs per person treated along with key details (e.g. country, number of participants, comparator, and cost components). Cost outcomes were further grouped by the costing perspective (i.e. only costs borne by healthcare providers, or societal costs which also include costs to patients and family members). For video-observed therapy (VOT) studies that only reported costs per treatment observation, costs were converted to a standard 6-month (26 week) treatment regimen with VOT performed 7X/week and DOT performed 5X/week to facilitate comparison: these were the most common observation frequencies for VOT and DOT respectively. For studies comparing VOT to DOT, we also present the cost per observation for all studies that reported scheduled observation frequencies.

When costs were available for multiple sites in a single country, only the weighted mean of all sites is presented, with weights reflecting the number of participants at each site. For such studies, information from individual sites is reported in supplement S4. Unless otherwise specified, reported costs for TB disease reflect drug-sensitive disease. Cost effectiveness results were grouped by outcome type (e.g. Cost per disability-adjusted life year [DALY] averted, cost per quality-adjusted life year [QALY] gained) and by costing perspective.

### Quality assessment

Quality of reporting was evaluated using the Consolidated Health Economic Evaluation Reporting Standards (CHEERS) 2022 checklist [17] for full text studies only. The checklist includes 28 criteria. A score of 1 was assigned for each criterion when fully met, 0 when it was not, and NA when it was not applicable. For each study, we calculated the percentage of checklist criteria met, after exclusion of those which were not applicable. This quality assessment was performed independently by two reviewers. Any differences between the two reviewers were resolved by consensus, with discussion with a third reviewer when necessary. Certainty of evidence was not rated formally but was discussed narratively.

### Patient and public involvement

Patients and the public were not specifically involved in the design, conduct, reporting, or dissemination plans of our research.

## RESULTS

### Search results

Of the 3,619 records identified by the initial search, 867 were removed as duplicates and 2,752 titles and abstracts were screened. Of these, 321 addressed DATs and TB and underwent full-text review for eligibility. 24 of these met inclusion criteria, while 5 others were identified by supplementary search of references and citations, for a total of 29 studies included in our review. Figure 1 shows the Preferred Reporting Items for Systematic Reviews and Meta-Analyses (PRISMA) 2020 flow chart for included and excluded studies.

**Figure 1:**
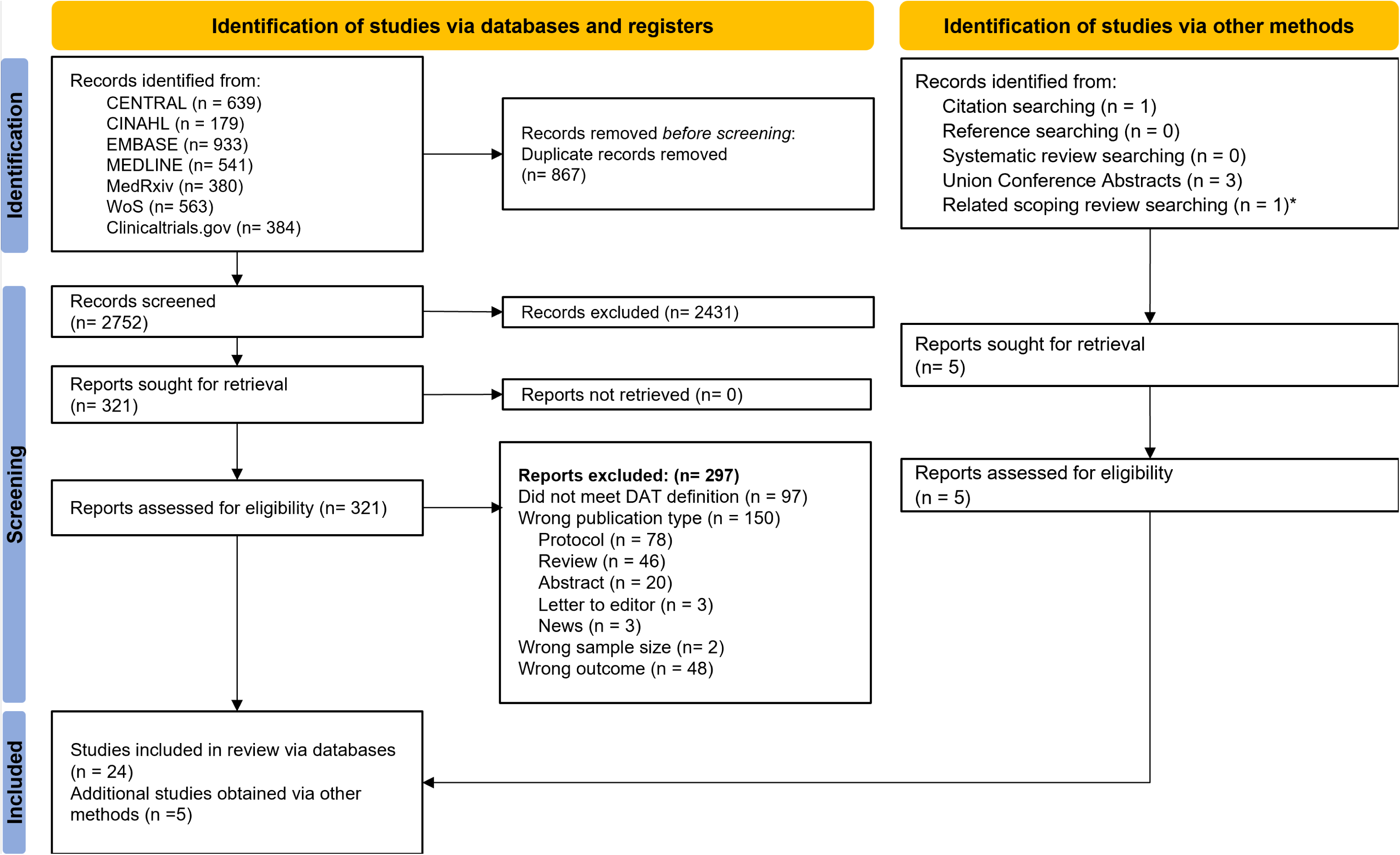
PRISMA flow diagram. *This study was captured by a related scoping review search by the authors of this systematic review and met all inclusion criteria

### Study characteristics

Detailed characteristics of each included study are listed in Table 1, while Figure 2 highlights the DAT types and country settings by income level. Over half the included studies evaluated video observed therapy (16 studies), followed by digital pillboxes (7 studies), SMS (4 studies) and medication sleeves with phone calls (“99DOTS”; 4 studies). There were 2 other DAT interventions addressed by one study each: automated interactive voice calls, and ingestible sensors.

**Figure 2:**
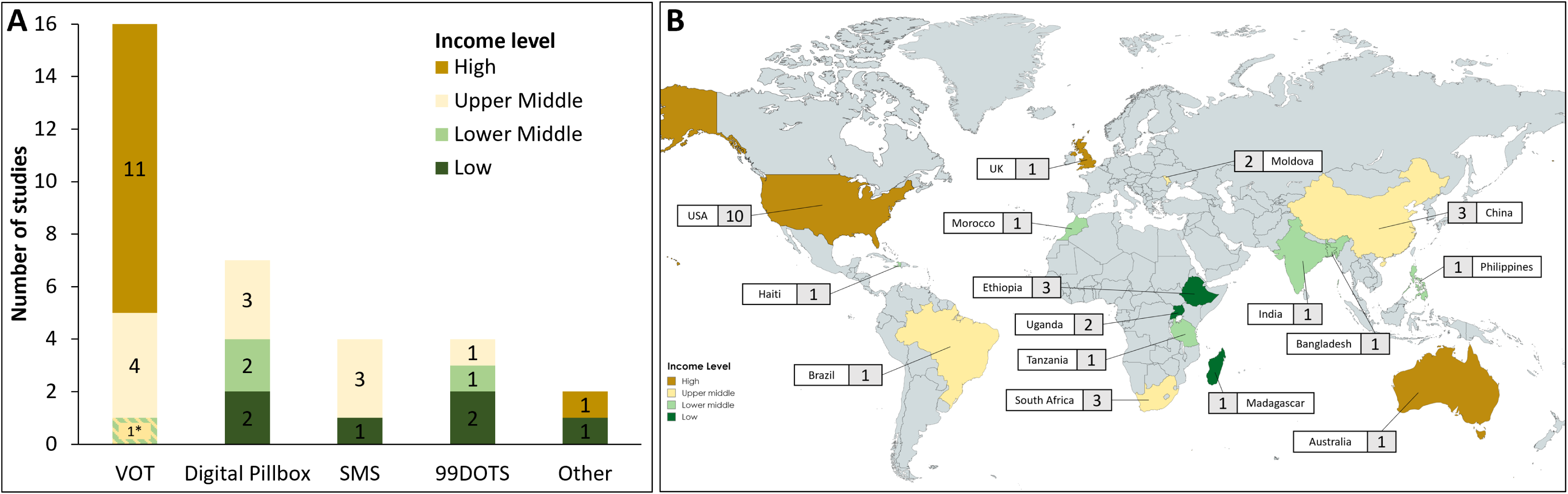
Summary of included studies. **A:** Number of studies evaluating each DAT type by country income level. *One study analyzed VOT in a lower middle and in upper middle income countries. Nsengiyumva et al. 2018 and Nsengiyumva et al. 2023 analyzed multiple DAT types (see Table 1) and thus are included in multiple columns above. **B:** Map of countries coloured by income level and number of included studies from each.

**Table 1:**
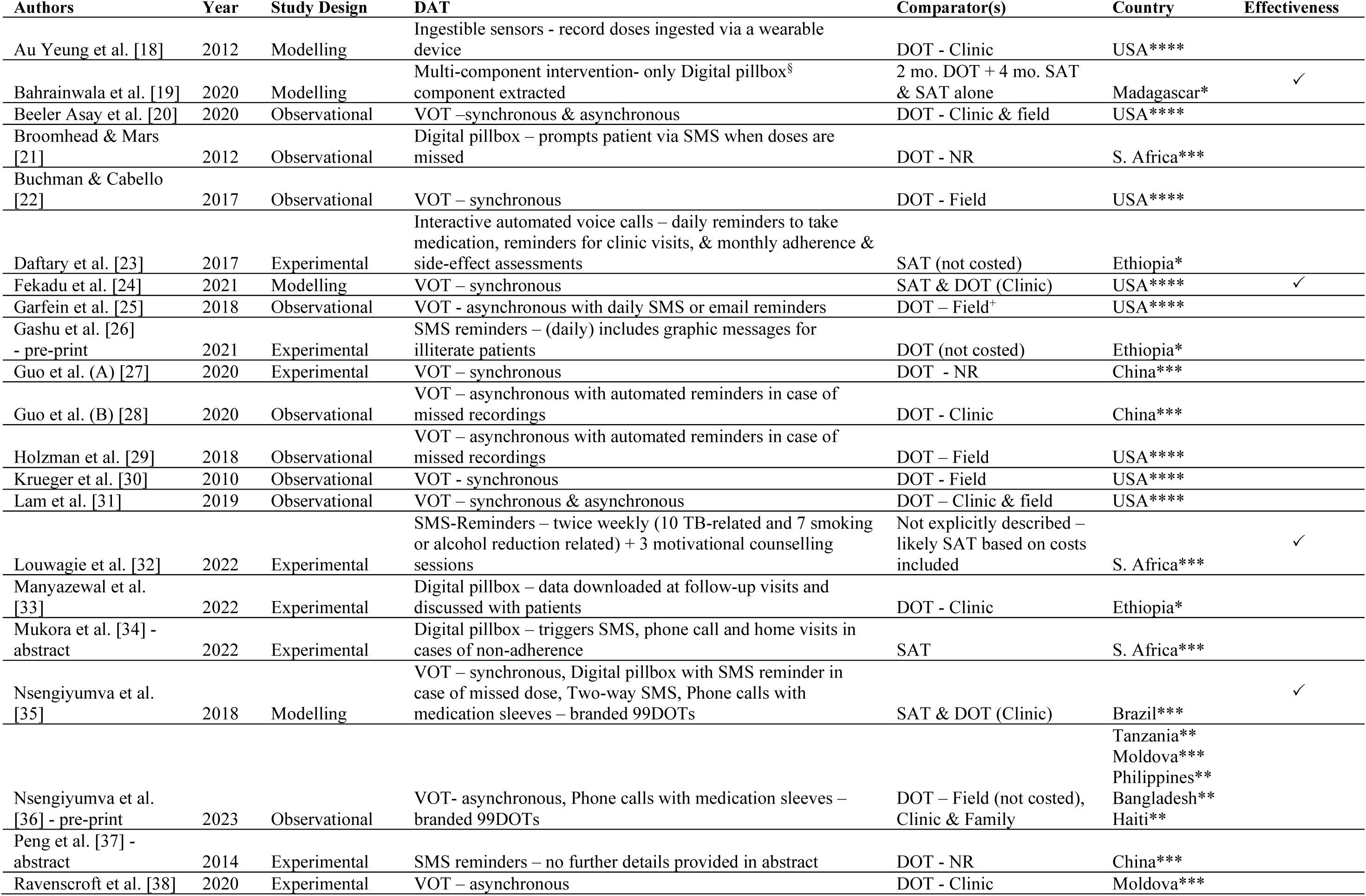
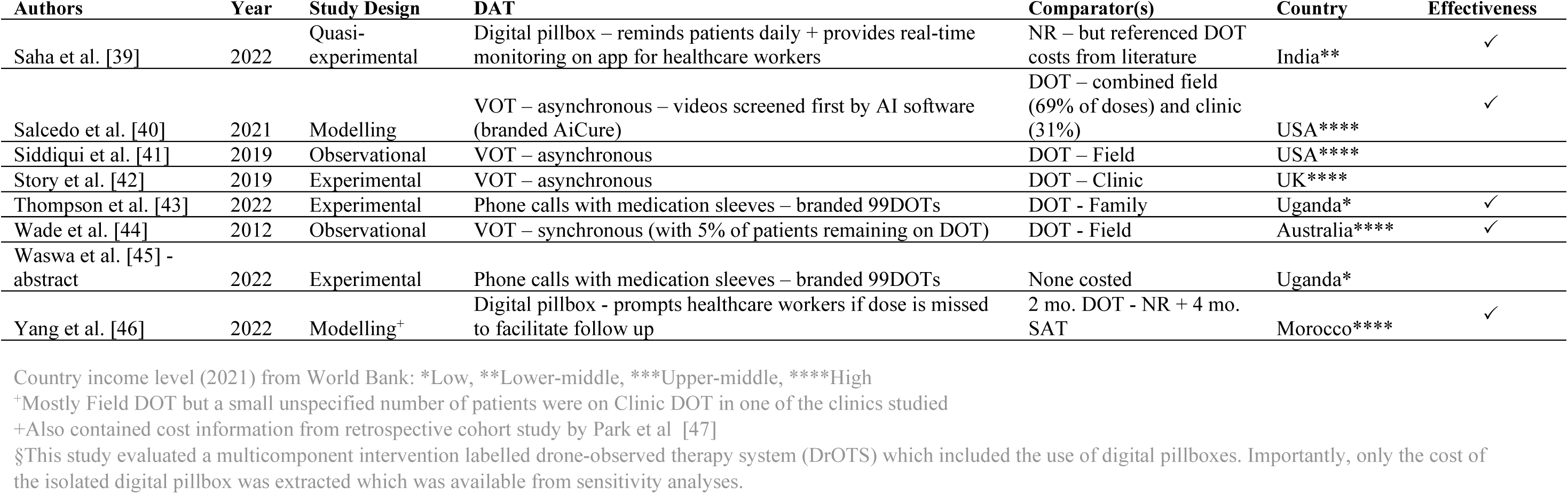
Characteristics of included studies.

**Table 2:**
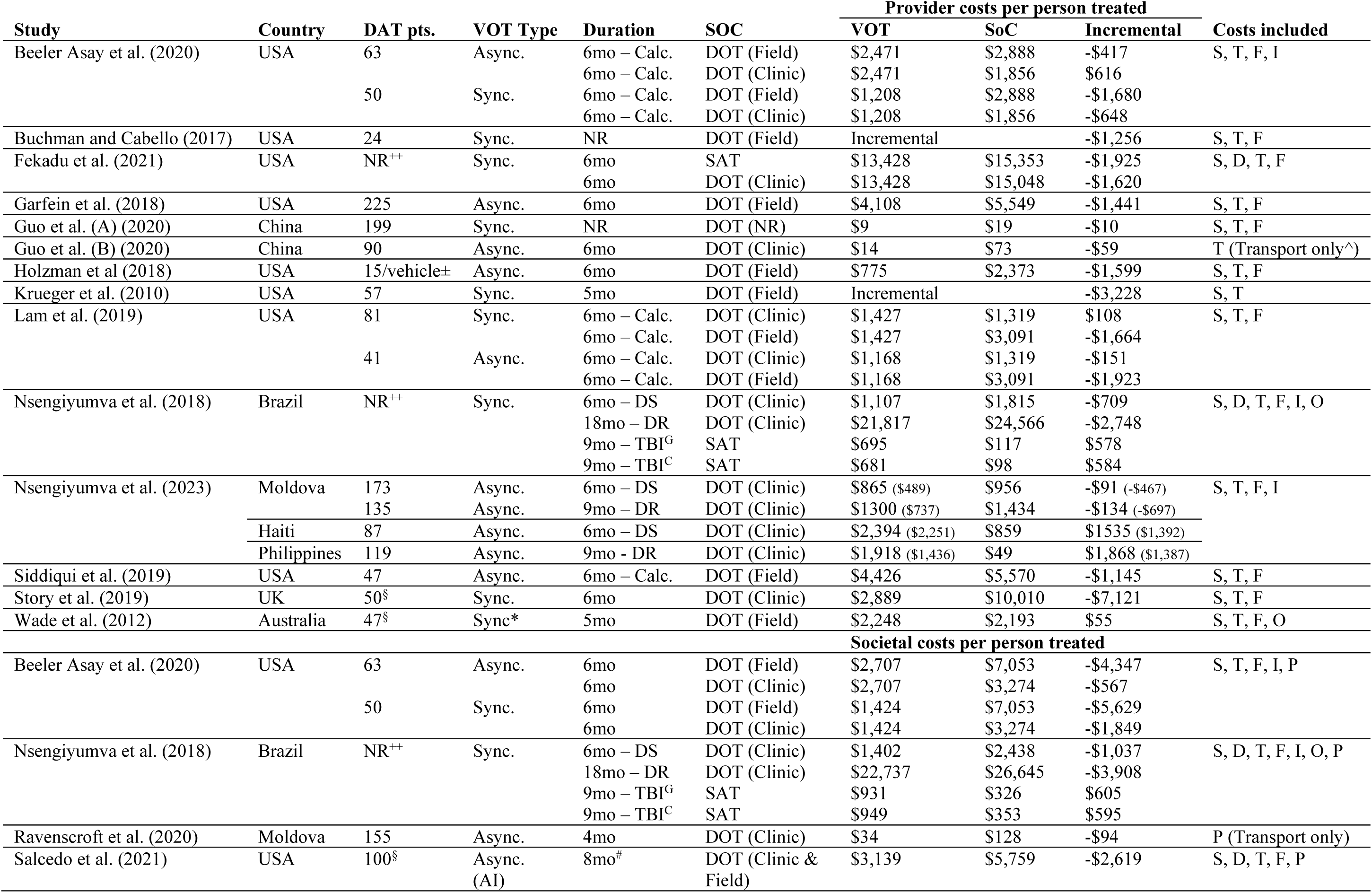
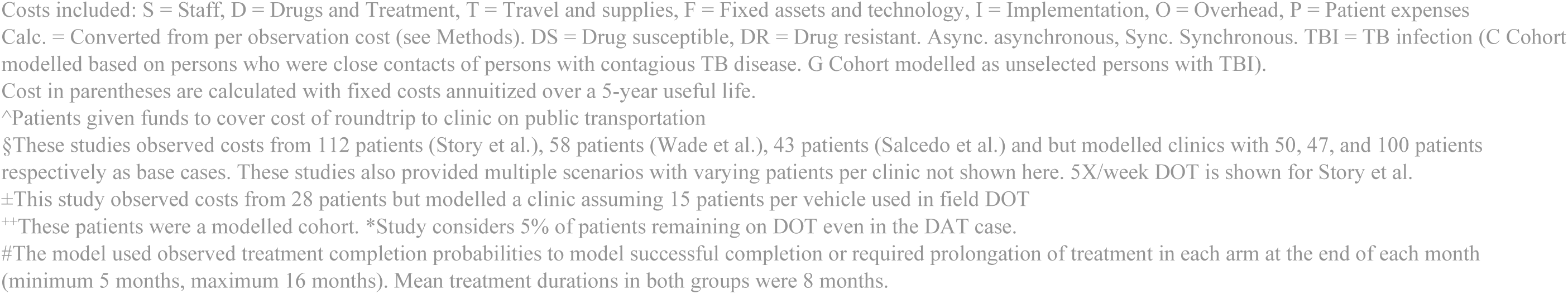
Provider and societal costs for VOT and standard of care (SoC)

The scope of included costs varied widely among the 26 full-text studies (Supplemental Table A2). For example, a few studies included up-front implementation costs, such as staff training and program set-up; most did not. Only six studies considered costs borne by persons with TB or their families, four of which also considered indirect costs such as lost wages. Eight studies included the cost of TB treatment, such as TB medication and follow-up testing. This was generally to model downstream cost savings associated with potential improvements in health outcomes. For example, if the intervention was thought to reduce acquired drug resistance, then the intervention would also reduce the additional treatment costs associated with multidrug resistant TB disease (MDR-TB). However, most studies did not consider such second-order effects.

### Video observed therapy (VOT)

Reported provider costs for VOT are summarized in Table 3. There was enormous variation, ranging from $9 per person treated in China, when only transport costs were considered [28] to $21,817 in a Brazil modeling study [35] which considered all costs including medications and follow-up testing for persons with MDR-TB. The median estimated provider cost across 15 reports was $1,364 per patient. For studies that reported costs for multiple sites in a single country, only the weighted average cost per patient is shown. The results for individual sites are provided in supplemental table A3. Three studies also considered scale-up scenarios [36,42,44] with larger hypothetical patient numbers that are not presented here. One analysis only considered averted DOT costs (such as fuel and labour) [30] but excluded any additional costs associated with VOT, so cost savings were inevitable and not necessarily reflective of all potential costs.

**Table 3:**
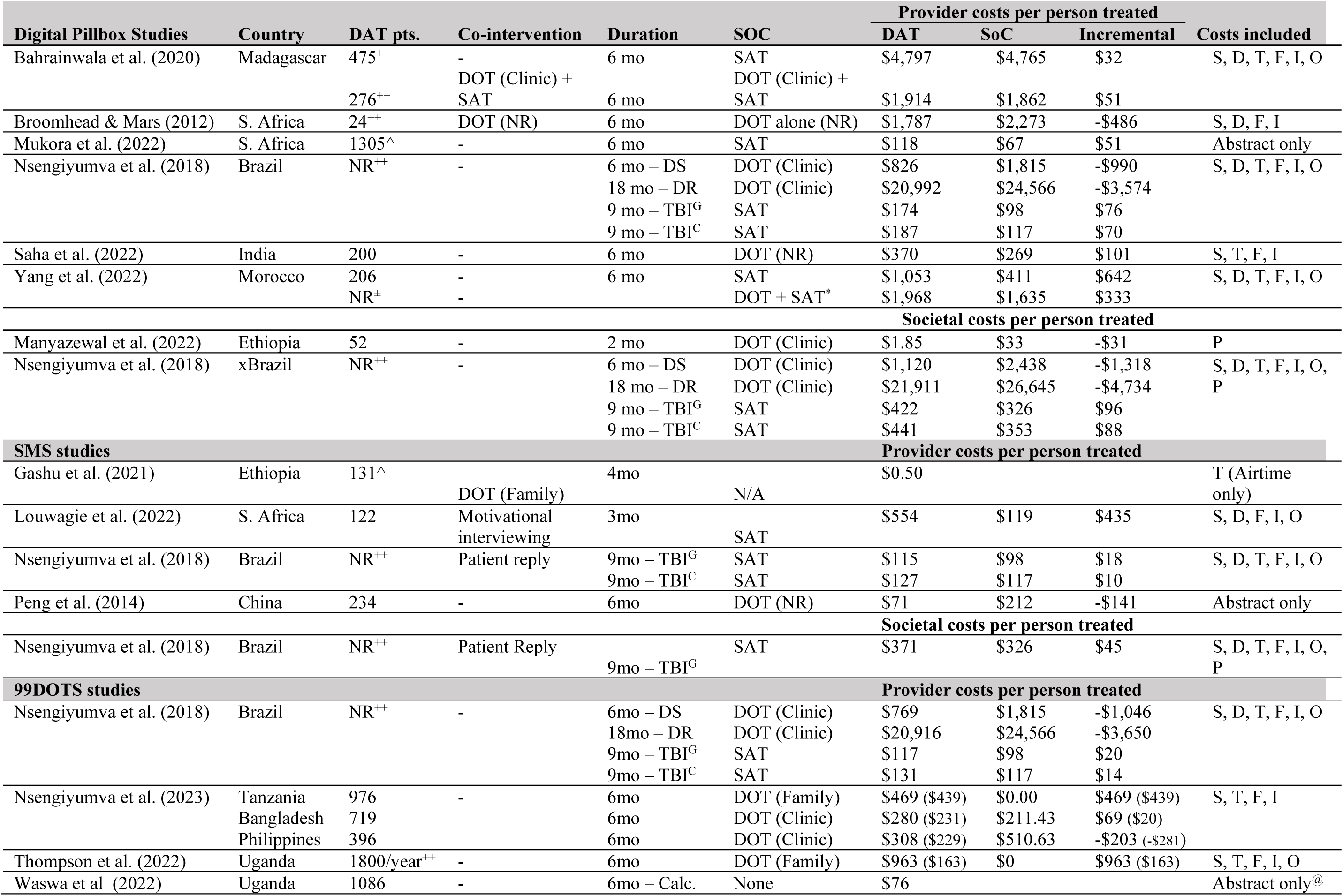
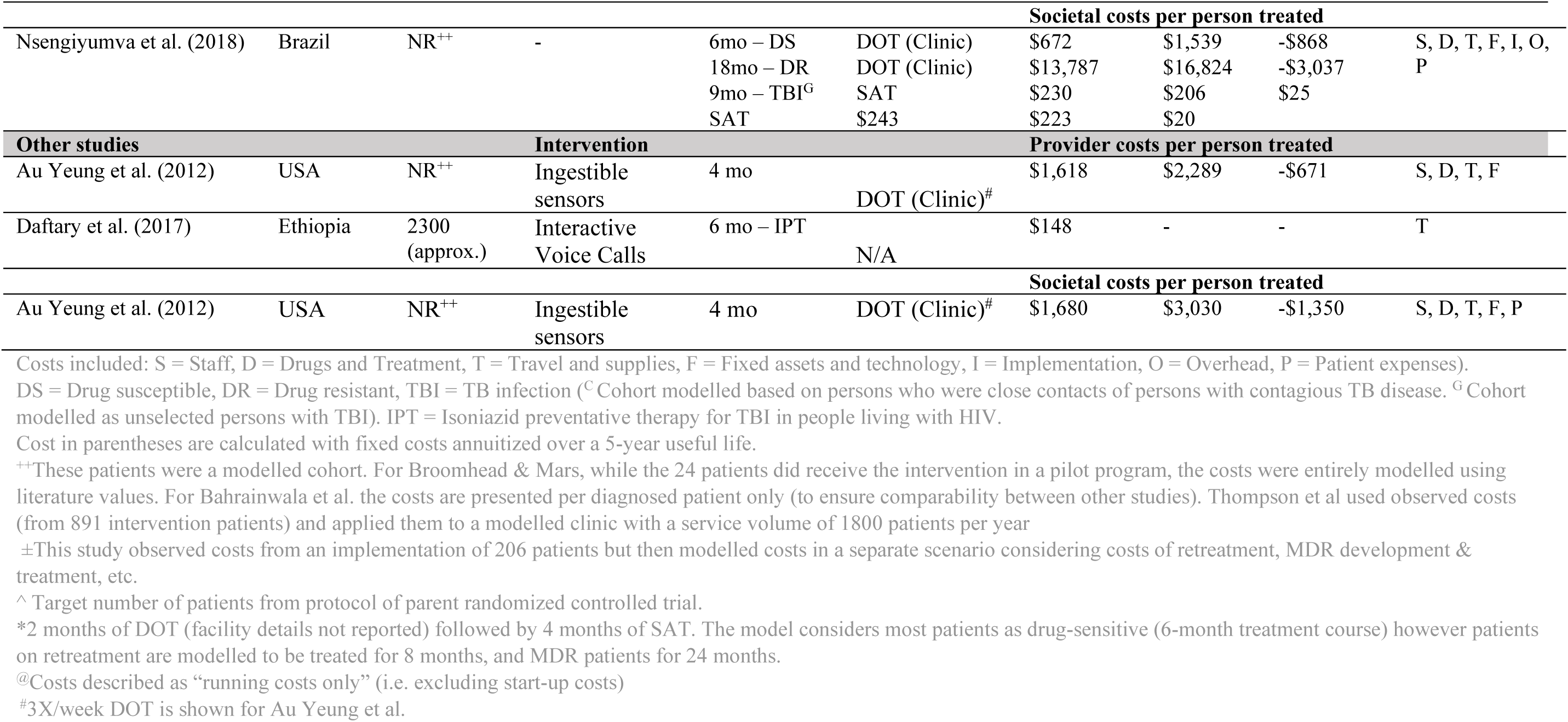
Provider and societal costs for DATs and the standard of care (SoC) grouped by type of intervention.

Most VOT studies took place in high-income (12 of 16) or upper middle-income (4 of 16) countries. Only one evaluated the use of VOT in a lower middle-income country [36]. Nine studies evaluated synchronous VOT while eight assessed asynchronous VOT, with both yielding savings in most scenarios.

In the two studies that also considered costs from the societal perspective, the analyses suggested cost savings to treated persons and their families with VOT, and hence further overall savings. A third study evaluated the use of an asynchronous VOT technology that leveraged artificial intelligence (AI) software (branded AiCure®) to screen the video recordings. From the societal perspective, VOT consistently yielded net savings over health care provider (field or clinic) DOT, but not compared to self-administered treatment. Two studies focused only on transportation costs reported by persons treated for TB; not surprisingly, VOT was then cheaper than clinic-based DOT [28,38]

A comparison of VOT and DOT costs per dose observation is shown in supplemental Table A4. Two VOT studies reported costs for both asynchronous and synchronous observation, compared in supplemental Table A5.

### Digital pillboxes

The provider costs for digital pillbox interventions are summarized in Table 3. The range of provider costs was again extremely wide (ranging from $118 to $20,992 per person) but generally similar to the standard of care which usually included at least some component of SAT. The median incremental cost for digital pillbox use was a 20% increase over the standard of care. While some studies evaluated digital pillboxes as a replacement for DOT, others considered it a means to augment DOT e.g., to support treatment of weekend doses which could not be directly observed. Only one study evaluated societal costs, and suggested savings relative to clinic-based DOT (Table 3). Another study that evaluated only patient expenses estimated that persons with TB who used a digital pillbox saved an average of $31 over the intensive phase of their treatment compared to the DOT group [33].

### SMS-based interventions

The provider costs for SMS-based interventions are summarized in Table 3. SMS was one of the lowest cost interventions at a median cost of $115 per person and were used exclusively in lower- and middle-income countries. The standard of care comparator was mostly SAT, except in one study that compared SMS with DOT (type unspecified) [37]; this was the only study that found SMS cost-saving. SMS based interventions were used in several clinical contexts including TBI treatment (9 months of isoniazid), throughout treatment of drug-sensitive TB disease, or for continuation phase treatment only. In one study, the intervention consisted of motivational interviewing along with SMS reminders [32]. The costs therefore include both the DAT and the interviewing co-intervention. While most other studies involved one-way SMS reminders, one report modelled a two-way system where patients confirmed whether the dose was taken by replying to the reminder message [35]. This study only evaluated the SMS intervention for persons treated for TBI. Only one study evaluated costs from a societal perspective (Table 3) and found similar costs when compared to self-administered therapy.

### Medication sleeves with phone-based dose recording (branded 99DOTS)

Four studies analyzed medication sleeve interventions in lower and middle-income countries, and the costs are summarized in Table 3. The provider costs for a 6-month regimen ranged from $76 in Uganda when excluding start-up costs [45] to $963 in another Uganda-based study that included those costs [43]. The intervention was associated with cost savings in most cases when compared against clinic DOT as standard of care. In the reports by Thompson et al and Nsengiyumva et al (2023; Tanzania group), there were no provider costs for DOT since the treatment observers were unpaid family members. The two studies also considered scenarios where fixed costs were annuitized over 5 years. Nsengiyumva et al. (2023) also considered scenarios for scale-up using larger theoretical patient numbers that are not presented here. Only one study [35] considered societal costs; it suggested additional savings for persons treated for TB disease.

### Other technologies (Interactive voice calls, and ingestible sensors)

One modelling study evaluated total treatment costs using a hypothetical target cost for ingestible sensors to detect and promote adherence to TB disease treatment [18]. This study modelled a clinic in a high-income country (USA) using ingestible sensors compared to clinic DOT as the standard of care. Another study provided feature phones and 6 months of airtime to support an interactive voice response intervention to promote adherence to TBI treatment [23]. The provider costs from these studies are highlighted in Table 3 along with results from the societal perspective reported by the ingestible sensors study.

A visual summary of all incremental costs to providers is illustrated in supplemental Figure A1. Some patterns become more evident visually. VOT was most often compared to some form of DOT, while other DATs were often compared to SAT. Not surprisingly, health provider savings with VOT were more pronounced when it was compared to field DOT, as opposed to facility-based DOT.

### Cost-effectiveness

Among the nine studies that addressed cost-effectiveness, outcomes assessed varied considerably. Four modeling studies estimated a cost per DALY averted; results are summarized in supplemental Table A6. Two modelling studies also estimated cost per TB case averted, largely among persons treated for TB infection (supplemental Table A7)

Three studies estimated cost per QALY gained (supplemental Table A8). One of these was a trial which found no significant effect of an SMS intervention plus motivational interviewing on health utility scores [32]. Hence the authors did not estimate cost per QALY gained; for completeness, we have done so using the point estimates they provided (supplemental Table A8). In a probabilistic sensitivity analysis, another study found that an AI-based VOT application [40] was dominant in 93.5% of simulations, i.e., cheaper with better health outcomes, compared to provider (clinic and field) DOT. In India, digital pillboxes were judged to be highly cost-effective compared to DOT [39].

One study compared synchronous VOT to field DOT and estimated it would cost $1.12 per additional treatment observation accomplished (95% confidence interval: $0.43-$1.91) to implement VOT in a clinic of 47 patients [44].

Another study compared an intervention involving medication sleeves and phone registration of doses (99DOTS) to family DOT in Uganda, and estimated it would cost $1,128 per additional treatment success (95% confidence interval: $697-$1,982) to implement the intervention in a clinic over 5 years [43]. Importantly however, this study used the per-protocol effect of this intervention, since the intention-to-treat analysis did not show improved treatment success with the DAT intervention in the parent randomized controlled trial.

### Subgroups and special populations

Few studies included persons treated for TB infection. Several combined data for persons treated for TB infection and disease, to estimate costs per observed treatment dose for their entire cohort [31, 20, 41] or presented estimates for a typical drug-sensitive treatment course [29]. One study reported on a group of persons living with HIV who received isoniazid preventive therapy for TB infection [23]. Only one study modelled costs and cost-effectiveness separately for persons treated for TBI, and found all DATs studied to be less cost-effective for TBI compared to active TB disease [35]. Of note, the standard of care was SAT for TBI and clinic-based DOT for active TB disease. This was also one of two studies that analyzed costs or cost-effectiveness separately for persons with drug-resistant TB disease; both studies suggested greater savings when DATs replaced clinic-based DOT for treatment support for drug-resistant TB, as compared to drug-sensitive disease [35,36]. Two studies modelled the development of acquired drug resistance among persons treated for TB disease, and included the ensuing treatment costs [21,46]. Several other studies combined data for persons with drug-sensitive and drug-resistant TB disease, with the latter group representing less than 5% of each cohort [20,29,38].

Only three studies included persons under the age of 16 [19,41], [36] and none performed any subgroup analysis according to age. As mentioned above, one study focused exclusively on isoniazid preventive therapy among persons living with HIV [23]. Two other cohorts included >10% persons living with HIV, but did not report any separate data for this subgroup [32,33]. Only two studies reported including any persons with extra-pulmonary TB disease, but again did not report data specific to this subgroup [38,42].

### International dollar vs. US dollar estimates

In lower- and middle-income settings, as expected the cost estimates in US dollars using market exchange rates (Supplemental Tables A9-A21) were 39-74% lower than those expressed in international dollars, using purchasing power parity. The difference was most marked for the 99DOTS medication sleeve intervention which was exclusively used in lower and middle-income settings and where the median provider cost of the intervention was 73% lower in US dollars. The median provider costs of digital pillbox and SMS interventions were 63% and 50% lower respectively. The cost-effectiveness estimates of these three interventions improved accordingly. As VOT cost estimates came from high-income countries, and those for the other DATs from lower- and middle-income country settings, the use of market exchange rates magnified cost differences between VOT and the lower-cost technologies.

### Quality of reporting

The results of the assessment for quality of reporting, using the CHEERS 2022 checklist [17], are summarized in Supplemental Table A9. The proportion of checklist items addressed ranged from 30% to 89% (median 68%). Five studies reported conflicts of interest, where a listed author was either an inventor of the DAT evaluated, or an employee of a company that manufactured it [18,25,29,39,44]. Two studies did not report on conflicts of interest [30,41].

## DISCUSSION

Of the technologies currently used by TB programs, video-observed therapy was reported as cost saving relative to clinic or field DOT in most studies, particularly with respect to cost per dose observed. However, VOT was primarily evaluated in high- or upper-middle income countries with well-established DOT programs, and technical infrastructure that can support VOT. It is unclear whether these results are generalizable to lower income settings where labor savings relative to in-person DOT may not offset equipment and infrastructure costs. Of course, from the provider perspective any treatment support provided by the health system, whether with a DAT or DOT, is more expensive than SAT.

In theory, costs for asynchronous VOT could differ from those for synchronous observation. Asynchronous VOT requires dedicated software and video storage but could allow further labour efficiencies if videos are watched consecutively or are screened by AI software. In fact, both asynchronous and synchronous VOT were reported to be cost-saving in higher-income settings. Indeed, the two studies that analysed both asynchronous and synchronous VOT generally found very similar costs per dose observed.

Digital pillbox costs were studied exclusively in lower- and middle-income countries, where they were occasionally used to supplement DOT. They were often judged cost saving when compared to DOT, particularly when costs to persons with TB were considered. However, they were not cost-effective or cost-saving when compared to SAT. Similarly, perhaps reflecting their low cost, SMS intervention costs were reported exclusively from lower and middle-income countries, and most often compared to SAT. However, interpretation was limited by significant co-interventions [32] or by restricted types of data available [26,37]. 99DOTS medication sleeves were also used primarily in lower- and middle-income countries, with estimated costs that were generally similar to or lower than those for clinic-based DOT.

The incremental cost of treatment support with DATs varied dramatically with the standard of care comparator and the setting. For example, savings to the health care system were typically greatest when DATs replaced field DOT (where the burden of travel is on the provider), as opposed to clinic DOT. There were also far more studies comparing DATs (particularly VOT) to DOT than to SAT. Given the considerable challenges in providing in-person DOT in lower- and middle-income countries [48–50], where most people with TB live, it is important to understand how the costs of the DATs compare to those for both DOT and SAT in the same settings.

The use of international dollars as opposed to USD from market exchange rates (reported in supplemental section S5) resulted in higher DAT and comparator costs for studies conducted in lower and middle-income countries. This is because purchasing power parity corrects for differences in resource prices between settings. Hence median costs for digital pillboxes, 99DOTS and SMS were 2-4 times higher in international dollars than in USD based on market exchange rates. The resulting cost estimates were therefore closer to (though still consistently lower than) those for VOT which was mostly used in the US. Cost effectiveness estimates from lower and middle-income countries showed a similar pattern.

Cost estimates based on international dollars and purchasing power parity are most useful for programs and decisionmakers considering adoption of DATs in their own settings. Estimates based on market exchange rates are more relevant to international funders and donors.

Overall, the quality of reporting was limited, when studies were assessed against the CHEERS checklist—a standard for health economic evaluations. For example, nearly half the studies we found did not report a currency year. Others incorrectly reported the perspective of their analyses, did not fully describe key data inputs, did not include relevant cost components, or contained apparent calculation errors.

### Strengths and limitations

To our knowledge, this is the first systematic review focusing specifically on cost and cost-effectiveness estimates for the application of digital adherence technologies to TB treatment. We used a wide-ranging search strategy across multiple databases, without language restriction and with inclusion of suitable publications from the grey literature—although the inclusion of conference abstracts limited the scope of information and quality assessment for those specific reports. Every step of our review, from filtering of titles and abstracts through data extraction, reporting, and quality assessment, involved two independent reviewers, with disagreements resolved by consensus, and a third senior reviewer when needed.

To support comparisons between study findings, we expressed all costs in the same currencies (2022 international dollars as well as 2022 US dollars). Similarly, we explicitly tabulated which cost components, such as technology and equipment, staff time, overheads, and patient/family costs were included in each study. Whenever possible, we summarized the cost of both the DAT and the comparator used for treatment support in each study setting. We also used a standard and widely used health economic analysis checklist to assess the quality of the reports included.

Because of the marked diversity of interventions, comparators, study settings, study designs, and cost measures, it was neither possible nor appropriate to pool study results through meta-analysis. For the cost-effectiveness analyses in particular, robust effectiveness data were often lacking. These analyses often involved the modeling of downstream costs and health outcomes, with many underlying (and unproven) assumptions. This was reflected in the diversity of results from these studies. Finally, there are likely other grey literature reports that were missed by our search.

## CONCLUSIONS

Cost and cost-effectiveness analyses for the digital adherence technologies currently used by TB programs have yielded variable results, particularly when compared to conventional directly observed treatment. Studies have often involved small numbers of affected persons, or specialized settings, hampering their generalizability. Video-observed treatment was more consistently associated with cost savings compared to clinic or field DOT in higher-income settings, related to reductions in travel and labor expenses for health care workers, and in productivity losses for persons on treatment. However, few analyses have considered costs borne by affected individuals and their families, so the overall potential for societal cost savings has not been adequately characterized. Any such savings are only relevant to the extent that treatment supported by DATs is associated with similar or better outcomes than the existing local standard of care. Moreover, any attendant health gains or cost savings can only be realized if the necessary technical and human resources are in place, and if barriers to DAT uptake are mitigated. It is also clear that more and higher quality operations research focusing on costs is needed, particularly in communities and settings that carry the greatest burden of TB.

## Supporting information

PRISMA checklist

## DECLARATIONS

## Acknowledgements

Dr. Jonathon R. Campbell provided invaluable technical expertise for economic analyses. Part of this work was presented in abstract and poster form at the 2023 North America Region Conference of the International Union of Tuberculosis and Lung Disease [51].

## Contributors

All authors contributed to the design of the research and approved the final manuscript. CK contributed to screening, data extraction and analysis, and drafted the manuscript. MSM contributed to the database search, screening, data extraction, and provided figures for the manuscript. MZ contributed to screening, and data extraction. GG contributed to the database search. CC, SB, and NF contributed to screening. KF, RS, and KS provided support in all steps of executing the review and revised the manuscript.

## Data availability statement

All data generated or analysed during this study are included in this published article and its supplementary information files.

## Changes to protocol

A sensitivity analysis with and without conference abstracts was not performed since only three abstracts were included and no meta-analysis was performed. Abstracts were instead clearly labelled on the results tables. A sensitivity analysis with market-based exchange rates was added. Certainty of evidence was not rated formally using criteria of the Grading of Recommendations, Assessment, Development and Evaluation (GRADE) approach since results were not pooled and cost-effectiveness outcomes were mostly not comparable.

