## Supplementary materials for "Cost and Cost-Effectiveness of Digital Technologies for Support of Tuberculosis Treatment Adherence: A Systematic Review"

Cedric Kafie [1], Mona Salaheldin Mohamed[1], Miranda Zary[1], Chimweta Ian Chilala [2], Shruti Bahukudumbi[3], Genevieve Gore[4], Nicola Foster[2], Katherine Fielding [2], Ramnath Subbaraman[3,5], and Kevin Schwartzman[1]

[1]McGill International Tuberculosis Centre; Research Institute of the McGill University Health Centre, Montréal, Canada
[2]TB Centre, London School of Hygiene and Tropical Medicine, London, UK
[3]Department of Public Health and Community Medicine, Tufts University School of Medicine, Boston, USA
[4]Schulich Library of Physical Sciences, Life Sciences, and Engineering; McGill University, Montréal, Canada
[5]Division of Geographic Medicine and Infectious Diseases, Tufts Medical Center, Boston, USA

### Data Supplement 1: Supplementary material

#### *S1 Search strategy*

##### MEDLINE (Ovid)

Ovid MEDLINE(R) ALL <1946 to April 18, 2023>

1 exp tuberculosis/ or mycobacterium tuberculosis/ 228099

2 Antitubercular Agents/ 41251

3 (Tuberculosis or Kochs disease or Phthisis or TB or MTB or MDRTB or XDRTB or DRTB or LTBI or directly observed treatment short course).mp. 300916

4 1 or 2 or 3 [TB CONCEPT] 306343

5 ((antitubercular agents/ or directly observed therapy/ or medication adherence/ or patient compliance/ or "treatment adherence and compliance"/) and technology/) or mobile applications/ or internet/ or cell phone/ or smartphone/ or text messaging/ or computer, handheld/ or telemedicine/ or therapy, computer-assisted/ or medical informatics applications/ 148201

6 (((digital* or electronic* or mobile or wireless* or virtual*) adj2 (adherence or medication monitor* or medication package* or observ*)) or digital technolog* or technology-based or Digital health or eHealth or e health or mhealth or m health or SMS or reminder* or short messag* service* or text messag* or MMS or multimedia messag* or MEMS or (monitor* adj2 (electronic* or sensor* or device*)) or Webcam* or web cam* or smartphone* or smart phone* or web based or health IT or health ICT or ((cell* or mobile) adj1 (device* or health or phone* or technolog*)) or video* or cellphone* or feature phone* or vdot or vmalt or tele* or dat or wot or 99dots or 99 dots or monitoring system* or ingestible sensor* or merm or artificial intelligence or ai or vot or ((digital or smart) adj (pill box* or pillbox*)) or VST).mp. 705421

7 5 or 6 [DAT CONCEPT] 772859

8 4 and 7 [TB CONCEPT AND DAT CONCEPT] 3219

9 limit 8 to yr="2000 -Current" 2791

10 Economics/ or exp "Costs and Cost Analysis"/ or Economics, Nursing/ or Economics, Medical/ or Economics, Pharmaceutical/ or exp Economics, Hospital/ or Economics, Dental/ or exp "Fees and Charges"/ or exp Budgets/ or exp models, economic/ or markov chains/ or monte carlo method/ or exp Decision Theory/ 394969

11 (Cost* or health payer* or societal or Economic* or Dominant or Dominated or Increment* or additional health gain* or Expense* or Budget* or Financi* or Benefit* or QALY*).mp. 2703781

12 (health utilit* or life year* or markov or monte carlo or pharmacoeconomic* or expenditure* or finance* or price or prices or pricing or (value adj2 (money or monetary)) or (decision* adj2 (tree* or analy* or model*))).mp.301385

13 or/10-12 [COST OR COST EFFECTIVENESS CONCEPT] 2875832

14 9 and 13 [TB CONCEPT AND DAT CONCEPT AND COST OR COST EFFECTIVENESS CONCEPT] 541

15 ("20220414" or "20220415" or "20220416" or "20220417" or "20220418" or "20220419" or 2022042* or 2022043* or 202205* or 202206* or 202207* or 202208* or 202209* or 20221* or 2023*).dt,ez,da. 1902583

16 14 and 15 82

https://ovidsp.ovid.com/ovidweb.cgi?T=JS&NEWS=N&PAGE=main&SHAREDSEARCHID=6Q0B8MrdSrEDSTHM9bRCUAwt58PjIrf3BXkSsDkuyGVifkVpHoqsrdglq7yGyHOJd

#### Embase (Ovid)

Embase <1996 to 2023 Week 15>

1 exp tuberculosis/ or mycobacterium tuberculosis/ 204337

2 tuberculostatic agent/ 30987

3 (Tuberculosis or Kochs disease or Phthisis or TB or MTB or MDRTB or XDRTB or DRTB or LTBI or directly observed treatment short course).mp. 246187

4 1 or 2 or 3 [TB CONCEPT] 257637

5 ((tuberculostatic agent/ or directly observed therapy/ or medication compliance/ or patient compliance/) and technology/) or communication technology/ or exp mobile application/ or internet/ or web-based intervention/ or exp mobile phone/ or text messaging/ or personal digital assistant/ or telemedicine/ or exp teleconsultation/ or telemonitoring/ or video consultation/ or computer-assisted therapy/ or computer-assisted drug therapy/ or medical informatics/ 261570

6 (((digital* or electronic* or mobile or wireless* or virtual*) adj2 (adherence or medication monitor* or medication package* or observ*)) or digital technolog* or technology-based or Digital health or eHealth or e health or mhealth or m health or SMS or reminder* or short messag* service* or text messag* or MMS or multimedia messag* or MEMS or (monitor* adj2 (electronic* or sensor* or device*)) or Webcam* or web cam* or smartphone* or smart phone* or web based or health IT or health ICT or ((cell* or mobile) adj1 (device* or health or phone* or technolog*)) or video* or cellphone* or feature phone* or vdot or vmalt or tele* or dat or wot or 99dots or 99 dots or monitoring system* or ingestible sensor* or merm or artificial intelligence or ai or vot or ((digital or smart) adj (pill box* or pillbox*)) or VST).mp. 870210

7 5 or 6 [DAT CONCEPT] 984663

8 4 and 7 [TB CONCEPT AND DAT CONCEPT] 5167

9 limit 8 to yr="2000 -Current" 5020

10 Economics/ or exp economic evaluation/ or exp *health economics/ or budget/ or exp economic model/ or exp markov chain/ or exp monte carlo method/ or Decision Theory/ 654528

11 (Cost* or health payer* or societal or Economic* or Dominant or Dominated or Increment* or additional health gain* or Expense* or Budget* or Financi* or Benefit* or QALY*).ti,ab,kf. 2903252

12 (health utilit* or life year* or markov or monte carlo or pharmacoeconomic* or expenditure* or finance* or price or prices or pricing or (value adj2 (money or monetary)) or (decision* adj2 (tree* or analy* or model*))).mp.434885

13 10 or 11 or 12 3379669

14 9 and 13 933

15 limit 14 to dc=20220414-20230419 131

https://ovidsp.ovid.com/ovidweb.cgi?T=JS&NEWS=N&PAGE=main&SHAREDSEARCHID=3DgeO1F774ECCcRfcqeXXAzvP09LOq5uzvf6SO0bpEKgOafK0X9ehQXyW219thL2h

#### CINAHL (EBSCOhost)

| # | Query | Limiters/Expanders | Last Run Via | Results |
|---|---|---|---|---|
| S16 | S14 AND S15 | Expanders - Apply equivalent subjects Search modes - Boolean/Phrase | Interface - EBSCOhost Research Databases Search Screen - Advanced Search Database - CINAHL Plus with Full Text | 39 |
| S15 | EM 20220414- OR ZD "in process" | Expanders - Apply equivalent subjects Search modes - Boolean/Phrase | Interface - EBSCOhost Research Databases Search Screen - Advanced Search Database - CINAHL Plus with Full Text | 1,147,669 |
| S14 | S9 AND S13 | Expanders - Apply equivalent subjects Search modes - Boolean/Phrase | Interface - EBSCOhost Research Databases Search Screen - Advanced Search Database - CINAHL Plus with Full Text | 179 |
| S13 | S10 OR S11 OR S12 | Expanders - Apply equivalent subjects Search modes - Boolean/Phrase | Interface - EBSCOhost Research Databases Search Screen - Advanced Search Database - CINAHL Plus with Full Text | Display |
| S12 | ("health utilit*" OR "life year*" OR markov OR "monte carlo" OR pharmacoeconomic* OR expenditure* OR finance* OR price OR prices OR pricing OR (value N2 (money OR monetary )) OR (decision* N2 (tree* OR analy* OR model* ))) | Expanders - Apply equivalent subjects Search modes - Boolean/Phrase | Interface - EBSCOhost Research Databases Search Screen - Advanced Search Database - CINAHL Plus with Full Text | Display |
| S11 | (Cost* OR "health payer*" OR societal OR Economic* OR Dominant OR Dominated OR Increment* OR "additional health gain*" OR Expense* OR Budget* OR Financi* OR Benefit* OR QALY*) | Expanders - Apply equivalent subjects Search modes - Boolean/Phrase | Interface - EBSCOhost Research Databases Search Screen - Advanced Search Database - CINAHL Plus with Full Text | Display |
| S10 | (MH "Economics") OR (MH "Costs and Cost Analysis+") OR (MH "Economic Aspects of Illness") OR (MH "Fees and Charges+") OR (MH "Budgets") OR (MH "Decision Trees+") | Expanders - Apply equivalent subjects Search modes - Boolean/Phrase | Interface - EBSCOhost Research Databases Search Screen - Advanced Search Database - CINAHL Plus with Full Text | Display |
| S9 | S4 AND S7 | Limiters - Published Date: 20000101-20231231 Expanders - | Interface - EBSCOhost Research Databases Search Screen - Advanced Search | 858 |

| | | | | |
|---|---|---|---|---|
| | | Apply equivalent subjects<br>Search modes - Boolean/Phrase | Database - CINAHL Plus with Full Text | |
| S8 | S4 AND S7 | Expanders - Apply equivalent subjects<br>Search modes - Boolean/Phrase | Interface - EBSCOhost Research Databases<br>Search Screen - Advanced Search<br>Database - CINAHL Plus with Full Text | Display |
| S7 | S5 OR S6 | Expanders - Apply equivalent subjects<br>Search modes - Boolean/Phrase | Interface - EBSCOhost Research Databases<br>Search Screen - Advanced Search<br>Database - CINAHL Plus with Full Text | Display |
| S6 | (((digital* OR electronic* OR mobile OR wireless* OR virtual*) N2 (adherence OR "medication monitor*" OR "medication package*" OR observ*)) OR "digital technolog*" OR technology-based OR "Digital health" OR eHealth OR "e health" OR mhealth OR "m health" OR SMS OR reminder* OR "short messag* service*" OR "text messag*" OR MMS OR "multimedia messag*" OR MEMS OR (monitor* N2 (electronic* OR sensor* OR device*)) OR Webcam* OR "web cam*" OR smartphone* OR "smart phone*" OR "web based" OR "health IT" OR "health ICT" OR ((cell* OR mobile) N1 (device* OR health OR phone* OR technolog*)) OR video* OR cellphone* OR "feature phone*" OR vdot OR vmalt OR tele* OR dat OR wot OR 99dots OR "99 dots" OR "monitoring system*" OR "ingestible sensor*" OR merm OR "artificial intelligence" OR ai OR vot OR ((digital OR smart) W1 ("pill box*" OR pillbox*)) OR VST) | Expanders - Apply equivalent subjects<br>Search modes - Boolean/Phrase | Interface - EBSCOhost Research Databases<br>Search Screen - Advanced Search<br>Database - CINAHL Plus with Full Text | Display |
| S5 | ( (MH "Antitubercular Agents") OR (MH "Directly Observed Therapy") OR (MH "Medication Compliance") OR (MH "Patient Compliance")) AND (MH "Technology") ) OR (MH "Wireless Communications") OR (MH "Mobile Applications") OR (MH "Internet") OR (MH "Internet-Based Intervention") OR (MH "Cellular Phone+") OR (MH "Text Messaging+") OR (MH "Instant Messaging") OR (MH "Interactive Voice Response Systems") OR (MH "Videoconferencing+") OR (MH "Computers, Hand-Held+") OR (MH "Telehealth+") OR (MH "Digital Technology") OR (MH "Therapy, Computer Assisted") OR (MH "Drug Therapy, Computer Assisted") OR (MH "Medical Informatics") | Expanders - Apply equivalent subjects<br>Search modes - Boolean/Phrase | Interface - EBSCOhost Research Databases<br>Search Screen - Advanced Search<br>Database - CINAHL Plus with Full Text | Display |
| S4 | S1 OR S2 OR S3 | Expanders - Apply equivalent subjects | Interface - EBSCOhost Research Databases<br>Search Screen - Advanced Search | Display |

| | | | | |
|---|---|---|---|---|
| | | Search modes - Boolean/Phrase | Database - CINAHL Plus with Full Text | |
| S3 | (Tuberculosis OR "Kochs disease" OR Phthisis OR TB OR MTB OR MDRTB OR XDRTB OR DRTB OR LTBI OR "directly observed treatment short course") | Expanders - Apply equivalent subjects Search modes - Boolean/Phrase | Interface - EBSCOhost Research Databases Search Screen - Advanced Search Database - CINAHL Plus with Full Text | Display |
| S2 | (MH "Antitubercular Agents") | Expanders - Apply equivalent subjects Search modes - Boolean/Phrase | Interface - EBSCOhost Research Databases Search Screen - Advanced Search Database - CINAHL Plus with Full Text | Display |
| S1 | (MH "Tuberculosis+") OR (MH "Mycobacterium Tuberculosis") | Expanders - Apply equivalent subjects Search modes - Boolean/Phrase | Interface - EBSCOhost Research Databases Search Screen - Advanced Search Database - CINAHL Plus with Full Text | Display |

#### CENTRAL (Cochrane Library/Wiley)

Search Name:

Date Run: 19/04/2023 19:42:11

Comment:

ID Search Hits

#1 (Tuberculosis or "Kochs disease" or Phthisis or TB or MTB or MDRTB or XDRTB or DRTB or LTBI or "directly observed treatment short course"):ti,ab,kw 9141

#2 (((digital* or electronic* or mobile or wireless* or virtual*) NEAR/2 (adherence or medication NEXT monitor* or medication NEXT package* or observ*)) or digital NEXT technolog* or technology-based or "Digital health" or eHealth or "e health" or mhealth or "m health" or SMS or reminder* or short NEXT messag* NEXT service* or text NEXT messag* or MMS or multimedia NEXT messag* or MEMS or (monitor* NEAR/2 (electronic* or sensor* or device*)) or Webcam* or web NEXT cam* or smartphone* or smart NEXT phone* or "web based" or "health IT" or "health ICT" or ((cell* or mobile) NEXT (device* or health or phone* or technolog*)) or video* or cellphone* or feature NEXT phone* or vdot or vmalt or tele* or dat or wot or 99dots or 99 NEXT dots or monitoring NEXT system* or ingestible NEXT sensor* or merm or artificial NEXT intelligence or ai or vot or ((digital or smart) NEXT (pill box* or pillbox*)) or VST):ti,ab,kw 359210

#3 #1 AND #2 with Publication Year from 2000 to 2023, in Trials 2118

#4 (Cost* or health NEXT payer* or societal or Economic* or Dominant or Dominated or Increment* or additional NEXT health NEXT gain* or Expense* or Budget* or Financi* or Benefit* or QALY*) 268651

#5 (health NEXT utilit* or life NEXT year* or markov or "monte carlo" or pharmacoeconomic* or expenditure* or finance* or price or prices or pricing or (value NEAR/2 (money or monetary)) or (decision* NEAR/2 (tree* or analy* or model*))):ti,ab,kw 23914

#6 #4 OR #5 277520

#7 #3 AND #6 639

Date added to CENTRAL trials database

Custom Range: 14/04/2022 to 19/04/2023

55 records

#### WOS.SCI,WOS.ISTP,WOS.ESCI (Web of Science Core Collection)

### Web of Science Search Strategy (v0.1)

Search: TS=(Tuberculosis OR "Kochs disease" OR Phthisis OR TB OR MTB OR MDRTB OR XDRTB OR DRTB OR LTBI OR "directly observed treatment short course" )

AND

TS=(((digital* OR electronic* OR mobile OR wireless* OR virtual* ) NEAR/2 (adherence OR "medication monitor*" OR "medication package*" OR observ* )) OR "digital technolog*" OR technology-based OR "Digital health" OR eHealth OR "e health" OR mhealth OR "m health" OR SMS OR reminder* OR "short messag* service*" OR "text messag*" OR MMS OR "multimedia messag*" OR MEMS OR (monitor* NEAR/2 (electronic* OR sensor* OR device* )) OR Webcam* OR "web cam*" OR smartphone* OR "smart phone*" OR "web based" OR "health IT" OR "health ICT" OR ((cell* OR mobile ) NEAR/1 (device* OR health OR phone* OR technolog* )) OR video* OR cellphone* OR "feature phone*" OR vdot OR vmalt OR tele* OR dat OR wot OR 99dots OR "99 dots" OR "monitoring system*" OR "ingestible sensor*" OR merm OR "artificial intelligence" OR ai OR vot OR ((digital OR smart ) NEAR/0 ("pill box*" OR pillbox* )) OR VST )

AND

(

TS=(Cost* OR "health payer*" OR societal OR Economic* OR Dominant OR Dominated OR Increment* OR "additional health gain*" OR Expense* OR Budget* OR Financi* OR Benefit* OR QALY*)

OR

TS=("health utilit*" OR "life year*" OR markov OR "monte carlo" OR pharmacoeconomic* OR expenditure* OR finance* OR price OR prices OR pricing OR (value N2 (money OR monetary )) OR (decision* N2 (tree* OR analy* OR model* )))

)

Editions: WOS.SCI,WOS.ISTP,WOS.ESCI

Timespan: 2000-01-01 to 2023-04-19

Date Run: Thu Apr 20 2023 16:05:49 GMT-0400 (Eastern Daylight Time)

Results: 563

### Database: Web of Science Core Collection

### Entitlements:

- WOS.IC: 1993 to 2023
- WOS.CCR: 1985 to 2023
- WOS.SCI: 1900 to 2023
- WOS.AHCI: 1975 to 2023
- WOS.BHCI: 2005 to 2023
- WOS.BSCI: 2005 to 2023
- WOS.ESCI: 2005 to 2023
- WOS.ISTP: 1990 to 2023
- WOS.SSCI: 1900 to 2023
- WOS.ISSHP: 1990 to 2023

### Searches:

Search:

#1

Timespan: 2022-04-14 to 2023-04-19

Date Run: Thu Apr 20 2023 16:07:32 GMT-0400 (Eastern Daylight Time)

Results: 70

#### MedRxiv and other preprints via Europe PMC

(TITLE:Tuberculosis OR TITLE:"Kochs disease" OR TITLE:Phthisis OR TITLE:TB OR TITLE:MTB OR TITLE:MDRTB OR TITLE:XDRTB OR TITLE:DRTB OR TITLE:LTBI OR TITLE:"directly observed treatment short course" OR TITLE:"antituberculosis" OR TITLE:"antituberculous") AND (Adher* OR "directly observed" OR Digital OR electronic OR internet OR mobile OR wireless OR virtual OR TITLE:technology OR "technology based" OR tele* OR ehealth OR "e health" OR mhealth OR "m health" OR SMS OR reminder* OR messaging OR message* OR MEMS OR web OR webcam* OR smartphone* OR "health IT" OR "health ICT" OR video* OR cellphone* OR phone* OR vdot OR vmalt OR dat OR wot OR 99dots OR "99 dots" OR "monitoring system" OR "monitoring systems" OR "ingestible sensor" OR "ingestible sensors" OR merm OR "artificial intelligence" OR AI OR VOT OR "smart pillbox" OR "smart pill box" OR VST OR "computer assisted") AND (SRC:PPR) AND CREATION_DATE:[2022-04-19 TO 2023-04-19]

#### Clinicaltrials.gov basic search

##### Translated and exported to EndNote/RIS and documented

384 records on April 25, 2023

384 Studies found for: **Adherence OR directly observed OR computer OR Digital OR electronic OR internet OR mobile OR virtual OR technology OR video OR mhealth OR artificial intelligence OR AI OR cellphones OR SMS OR reminders OR monitoring OR MEMS OR DAT OR sensors | Tuberculosis OR Kochs disease OR Phthisis OR TB OR MTB OR MDRTB OR XDRTB OR DRTB OR LTBI OR directly observed treatment short course**

*S2  Definition for inclusion of digital adherence technologies*

Intervention can be patient-facing only, provider-facing only, or patient and provider-facing.

Intervention includes:

- a digital component (which could be part of a multi-component intervention) that goes beyond a means of non-automated voice or text communication between a human operator and a patient (i.e. more than a manual phone call, text message or email)
- with the intention to promote treatment adherence and/or reducing missed visits and/or reducing LTFU (and thereby improving successful treatment outcomes)

*Examples of "digital adherence component" included (though not limited to)*

- SMS daily reminders (1- or 2-way) to patient to take treatment
- SMS reminders (1- or 2-way) to patient to attend dispensing visit/routine follow-up appointment
- Automated phone calls to remind patient for visit/daily dose
- Smart pillbox daily reminders to patient to take treatment
- Smart pillbox reminders to patient attend dispensing visit/routine follow-up appointment
- Chatbot/telemedicine accessed by TB patients that provides information about "treatment adherence"
- Automated feedback (such as electronic health record)  to alert HCW to a missed visit (eg., dispensing, routine follow-up appt for patients on TB/LTBI treatment) by a patient with an intended action of "promoting treatment adherence and/or reducing LTFU"
- Digital calendar generated (from electronic health record/smart pill box) showing missed visits/doses by patient at consultation with HCW
- Telehealth – video-call initiated by HCW
- WhatsApp group (with patients/HCWs)-  educational messages sent etc..
- Specialist Healthcare provider Apps (– not automated) – used by patient and HCW

These need to be used *in conjunction* with the intention of promoting treatment adherence and/or reducing missed visits and/or reducing LTFU (and thereby improving successful treatment outcomes)

*Examples of excluded interventions (not limited to)*

- Electronic health record to document (monitor/record/summarise) visit attendance only - with NO intended action of "promote treatment adherence and/or reducing LTFU"
- Mobile technology to collect data (TB register etc..) with no feedback loop to promote adherence
- Non-automated "routine telephone calls" to patient
- Phone calls or SMS or WhatsApp messages that are not automated (i.e. written and sent by a human) - with no other digital component

#### *S3 Cost component checklist*

*Table A1: Cost component checklist*

| Category | Item |
|---|---|
| Staff | Healthcare worker salary & benefits |
| TB treatment | TB treatment costs (drugs, follow-up lab tests) |
| Travel and Supplies | DAT-specific supplies (if applicable, e.g. phone number medication sleeves) |
| | Technology supplied to patients that is not recouped (e.g. given a phone, tablet, wireless sensor) |
| | Travel costs (during operation, if applicable) |
| | Airtime, data, and other variable fees |
| Fixed assets & technology | DAT technology support (& software-as-a-service fees) |
| | DAT components (e.g. computers, projectors, smartphones, software, items in the clinic) |
| | DAT technology servers (if applicable) |
| Design & Implementation | DAT design and production |
| | Training costs - salaries of health workers and the trainers, lodging, travel, training materials |
| | Travel costs (during implementation) |
| Overhead | Rent (or building depreciation) |
| | Utilities |
| | General administrative supplies |
| Other provider costs | Additional costs not captured above - Health Care System |
| Out-of-pocket patient costs | Out-of-pocket healthcare costs (drugs, fees) |
| | Travel costs |
| | Data & airtime expenses |
| | Technology purchased by patients |
| Indirect patient costs | Lost productivity (e.g. time off work) |
| Other patient costs | Additional costs not captured above - Patients / Family |

#### *S4 Supplemental results*

*Table A2: Summary of included costs in each full text study*

| | | Provider costs | | | | | | Patient Costs | |
|---|---|---|---|---|---|---|---|---|---|
| **Study** | **DATs assessed** | **Staff** | **TB drugs & treatment** | **Travel & supplies** | **Fixed assets & technology** | **Implement-ation** | **Overhead** | **Out-of-pocket** | **Indirect** |
| Bahrainwala et al. (2020) | Digital Pillboxes | ☑ | ☑ | ✓ | ☑ | ✓ | ☑ | | |
| Broomhead & Mars (2012) | Digital Pillboxes | ☑ | ☑ | | ☑ | ✓ | | | |
| Manyazewal et al. (2022) | Digital Pillboxes | | | | | | | ☑ | ☑ |
| Yang et al (2022) | Digital Pillboxes | ☑ | ☑ | ☑ | ☑ | ✓ | ☑ | | |
| Saha et al. (2022) | Digital Pillboxes | ☑ | | ☑ | ☑ | ☑ | | | |
| Au Yeung et al. (2012) | Ingestible Sensors | ☑ | ☑ | ✓ | ☑ | | | ✓ | ☑ |
| Nsengiyumva et al. (2018) | Multiple | ☑ | ☑ | ☑ | ✓ | ✓ | ✓ | ☑ | ☑ |
| Nsengiyumva et al. (2023) | Multiple | ☑ | | ☑ | ☑ | ☑ | | | |
| Daftary et al. (2017) | Phone-based | | | ☑ | | | | | |
| Thompson et al. (2022) | Phone (Pill sleeve) | ☑ | | ☑ | ☑ | ☑ | ☑ | | |
| Gashu et al. (2021) | SMS-Based Only | | | ✓ | | | | | |
| Louwagie et al. (2022) | SMS-Based Only | ☑ | ☑ | | ✓ | ☑ | ✓ | | |
| Beeler Asay et al. (2020) | VOT | ☑ | | ☑ | ✓ | ✓ | | ☑ | ☑ |
| Buchman & Cabello (2017) | VOT | ☑ | | ☑ | ✓ | | | | |
| Fekadu et al. (2021) | VOT | ☑ | ☑ | ✓ | ✓ | | | | |
| Garfein et al. (2018) | VOT | ☑ | | ☑ | ✓ | | | | |
| Guo et al. (A) (2020) | VOT | Unclear | | ✓ | ✓ | | | | |
| Guo et al. (B) (2020) | VOT | | | ✓ | | | | | |
| Holzman et al (2018) | VOT | ☑ | | ☑ | ☑ | | | | |
| Krueger et al. (2010) | VOT | ☑ | | ✓ | | | | | |
| Lam et al. (2019) | VOT | ☑ | | ☑ | ✓ | | | | |
| Ravenscroft et al. (2020) | VOT | | | | | | | ✓ | |
| Salcedo et al. (2021) | VOT | ☑ | ☑ | ☑ | ☑ | | | ☑ | |
| Siddiqui et al. (2019) | VOT | ☑ | | ☑ | ✓ | | | | |
| Story et al. (2019) | VOT | ☑ | | ☑ | ☑ | | | | |
| Wade et al. (2012) | VOT | ☑ | | ☑ | ☑ | | ☑ | | |
| **Total studies considering category:** | | **20/26** | **8/26** | **22/26** | **20/26** | **9/26** | **6/26** | **6/26** | **4/26** |

S5 ☑Category included in costs. ✓Only one item from category included in costs.

*Table A3: Provider costs for VOT and standard of care (SoC) for studies with multiple sites (see* ***Error! Reference source not found.****2 in main text)*

| Study | Country | Site | DAT pts. | VOT Type | Duration | SOC | Provider costs per patient VOT | Provider costs per patient SoC | Provider costs per patient Incremental | Costs included |
|---|---|---|---|---|---|---|---|---|---|---|
| Beeler Asay et al. (2020) | USA | NYC | 186/year | Async. | 6mo – Calc. | DOT (Field) | $1,394 | $3,375 | -$1,980 | S, T, F, I |
| | | NYC | 186/year | Async. | 6mo – Calc. | DOT (Clinic) | $1,394 | $1,054 | $340 | |
| | | NYC | 399/year | Sync. | 6mo – Calc. | DOT (Field) | $1,208 | $3,375 | -$2,167 | |
| | | NYC | 399/year | Sync. | 6mo – Calc. | DOT (Clinic) | $1,208 | $1,054 | $154 | |
| | | RI | 9/year | Async. | 6mo – Calc. | DOT (Field) | $4,624 | $3,139 | $1,485 | |
| | | SF | 62/year | Async. | 6mo – Calc. | DOT (Field) | $1,491 | $2,449 | -$959 | |
| | | SF | 62/year | Async. | 6mo – Calc. | DOT (Clinic) | $1,491 | $3,942 | -$2,452 | |
| Krueger et al. (2010) | USA | Pierce | 41 | Sync.* | 5mo | DOT (Field) | Incremental only | | -$3,171 | S, T |
| | | Squamish | 16 | Sync.* | 5mo | DOT (Field) | Incremental only | | -$3,373 | |

Note: Beeler Asay et al. did not provide a specific site breakdown on the number of patients that made up the combined sample of 63 patients for asynchronous and 57 patients for synchronous VOT. They did provide however annual patient volumes for each site.
*Study considers savings in patients that started on DOT and were switched to VOT for variable portions of their treatment (average of 20 weeks on VOT for all patients)

Beeler Asay et al. had 3 study sites in the United States described as New York City, Rhode Island and San Francisco. The Rhode Island site had a much smaller case load than the other sites and so fixed software costs for asynchronous VOT were proportionally much higher and represented over 80% of the per patient cost. Societal costs were not broken down by site for this study. While costs were presented in the study on a per session basis, the costs were converted to per patient using the process described in *Methods*.

*Table A4: Provider costs for VOT and DOT on a per observation basis*

| | | | | | Provider costs per observation | | | |
|---|---|---|---|---|---|---|---|---|
| **Study** | **Country** | **DAT pts.** | **VOT Type** | **SOC** | **VOT** | **SoC** | **Incremental** | **Costs included** |
| Beeler Asay et al. (2020) | USA | 63 | Async. | DOT (Field) | $13.58 | $22.22 | -$8.64 | S, T, F, I |
| | | | | DOT (Clinic) | $13.58 | $14.27 | -$0.69 | |
| | | 50 | Sync. | DOT (Field) | $6.64 | $22.22 | -$15.58 | |
| | | | | DOT (Clinic) | $6.64 | $14.27 | -$7.64 | |
| Fekadu et al. (2021) | USA | NR[++] | Sync. | DOT (Clinic) | $79.93 | $125.40 | -$45.47 | S, D, T, F |
| Garfein et al. (2018) | USA | 225 | Async. | DOT (Field) | $22.57 | $42.68 | -$20.11 | S, T, F |
| Guo et al. (B) (2020) | China | 90 | Async. | DOT (Clinic) | $0.14 | $1.05 | -$0.91 | T (Transport only^) |
| Holzman et al (2018) | USA | 15/vehicle[±] | Async. | DOT (Field) | $5.96 | $18.26 | -$12.30 | S, T, F |
| Krueger et al. (2010) | USA | 57 | Sync. | DOT (Field) | Incremental | | -$41.62 | S, T |
| Lam et al. (2019) | USA | 81 | Sync. | DOT (Clinic) | $7.84 | $10.14 | -$2.30 | S, T, F |
| | | | | DOT (Field) | $7.84 | $23.78 | -$15.94 | |
| | | 41 | Async. | DOT (Clinic) | $6.41 | $10.14 | -$3.73 | |
| | | | | DOT (Field) | $6.41 | $23.78 | -$17.36 | |
| Nsengiyumva et al. (2018) | Brazil | NR[++] – DS | Sync. | DOT (Clinic) | $14.19 | $23.27 | -$9.08 | S, D, T, F, I, O |
| | | NR[++] – DR | Sync. | DOT (Clinic) | $93.24 | $104.98 | -$11.75 | |
| Nsengiyumva et al. (2023) | Moldova | 173 – DS | Async. | DOT (Clinic) | $4.81 ($2.72) | $7.97 | -$3.16 (-$5.25) | S, T, F, I |
| | | 135 – DR | Async. | DOT (Clinic) | $4.81 ($2.72) | $7.97 | -$3.16 (-$5.25) | |
| | Haiti | 87 | Async. | DOT (Clinic) | $13.30 ($12.51) | $7.16 | $6.14 ($5.35) | |
| | Philippines | 119 | Async. | DOT (Clinic) | $7.10 ($5.32) | $0.27 | $6.83 ($5.05) | |
| Siddiqui et al. (2019) | USA | 47 | Async. | DOT (Field) | $24.32 | $42.85 | -$18.53 | S, T, F |
| Story et al. (2019) | UK | 50[§] | Sync. | DOT (Clinic) | $15.87 | $77.00 | -$61.13 | S, T, F |
| Wade et al. (2012) | Australia | 47[§] | Sync** | DOT (Field) | $15.94 | $23.84 | -$7.89 | S, T, F, O |

Costs included: S = Staff, D = Drugs and Treatment, T = Travel and supplies, F = Fixed assets and technology, I = Implementation, O = Overhead, P = Patient expenses

DS = Drug susceptible, DR = Drug resistant. Cost in parentheses are calculated with fixed costs annuitized over a 5-year useful life.

§These studies observed costs from 112 patients (Story et al.) and 58 patients (Wade et al.) but modelled clinics with 50 and 47 patients respectively as base cases. These studies also provided multiple scenarios with varying patients per clinic not shown here. 5X/week DOT is shown for Story et al.

^Patients given funds to cover cost of roundtrip to clinic on public transportation

±This study observed costs from 28 patients but modelled a clinic assuming 15 patients per vehicle used in field DOT. [++]These patients were a modelled cohort.

**Study considers 5% of patients remaining on DOT even in the DAT case

*Table A5:* Provider costs for synchronous and asynchronous VOT on a per observation basis

| Study | Country | Async. Pts. | Sync. Pts. | Setting | Provider costs per observation | | | Costs Included |
|---|---|---|---|---|---|---|---|---|
| | | | | | Asynchronous | Synchronous | Incremental | |
| Lam et al. (2019) | USA | 81 | 41 | **New York City** | **$6.41** | **$7.84** | **-$1.43** | S, T, F |
| Beeler Asay et al. (2020) | USA | 257/year | 399/year | All sites | $13.58 | $6.64 | $6.94 | S, T, F, I |
| | | 186/year | 399/year | **New York City** | **$7.66** | **$6.64** | **$1.02** | |
| | | 62/year | - | San Francisco | $8.19 | - | | |
| | | 9/year | - | Rhode Island | $25.40 | - | | |

Costs included: S = Staff, D = Drugs and Treatment, T = Travel and supplies, F = Fixed assets and technology, I = Implementation, O = Overhead, P = Patient expenses
Note: Beeler Asay et al. did not report a site breakdown on the number of patients that made up the combined sample of 63 patients for asynchronous and 57 patients for synchronous VOT. They provided annual patient volumes for each site.

*Table A6: Incremental cost per DALY averted from provider and societal perspectives*

| Study | Country | DAT pts. | DAT used | Intervention Duration | SoC | ICER - $/DALY averted # Health system | ICER - $/DALY averted # Societal | Costs included |
|---|---|---|---|---|---|---|---|---|
| Bahrainwala et al. (2020) | Madagascar | 276⁺⁺ | Digital pillbox | 6mo | DOT | $124§ | | S, D, T, F, I, O |
| | | 445⁺⁺ | | 6mo | SAT | $76§ | | |
| Fekadu et al. (2021) | United States | NR⁺⁺ | VOT | 6mo | DOT | DAT Dominant§ | | S, D, T, F |
| | | NR⁺⁺ | | 6mo | SAT | DAT Dominant& | | |
| Nsengiyumva et al. (2018) | Brazil | NR⁺⁺ | 99DOTS | 9mo – TBI$^C$ | SAT | $410 ($20 to $869) | $1,029 (-$195 to $2647) | S, D, T, F, I, O, (P) |
| | | NR⁺⁺ | | 9mo – TBI$^G$ | SAT | $2,674 ($1247 to $3609) | $5,721 ($1569 to $10752) | |
| | | NR⁺⁺ | SMS + reply | 9mo – TBI$^C$ | SAT | $240 (-$111 to $607) | $859 (-$328 to $2,387) | |
| | | NR⁺⁺ | | 9mo – TBI$^G$ | SAT | $1,952 ($748 to $2,887) | $4,999 ($1,044 to $10,084) | |
| | | NR⁺⁺ | VOT (sync.) | 9mo – TBI$^C$ | SAT | $19,139 ($7,140 to $34,347) | $19,758 ($7,281 to $34,252) | |
| | | NR⁺⁺ | | 9mo – TBI$^G$ | SAT | $82,923 ($40,082 to $118,490) | $85,970 ($40,965 to $121,906) | |
| | | NR⁺⁺ | Digital pillbox | 9mo – TBI$^C$ | SAT | $2,299 ($732 to $4,238) | $2,918 ($759 to $5,468) | |
| | | NR⁺⁺ | | 9mo – TBI$^G$ | SAT | $10,775 ($5,313 to $15,261) | $13,822 ($6,012 to $20,418) | |
| Yang et al. (2022) | Morocco | NR⁺⁺ | Digital pillbox | 6mo* | DOT+SAT | $1,146&& | | S, D, T, F, I, O |

Costs included: S = Staff, D = Drugs and Treatment, T = Travel and supplies, F = Fixed assets and technology, I = Implementation, O = Overhead, P = Patient expenses

TBI = TB infection ($^C$ Cohort modelled based on persons who were close contacts of persons with contagious TB disease. $^G$ Cohort modelled as unselected persons with TBI).

⁺⁺These patients were a modelled cohort. #Negative values indicate cost savings. Values in parentheses are 95% uncertainty intervals

*The model considers most patients as drug-sensitive (6 months) however patients on retreatment are modelled to be treated for 8 months, and MDR patients for 24 months

§Calculated from available data in sensitivity analyses and therefore no uncertainty range available

&95% uncertainty range via probabilistic sensitivity analysis: Reduction in DALYs (0.4385-0.0.4440) and reduction in cost ($1,924-$2,082)

&&95% uncertainty range via probabilistic sensitivity analysis: Reduction in DALYs (0.08-0.32), increased cost ($246-417)

*Table A7: Incremental cost per TB case averted with provider and societal perspectives.*

| Study | Country | DAT pts. | DAT used | Intervention Duration | SoC | ICER - $/case averted# Provider | ICER - $/case averted# Societal | Costs included |
|---|---|---|---|---|---|---|---|---|
| Nsengiyumva et al. (2018) | Brazil | NR++ | 99DOTS | 9mo – TBI^C | SAT | $2,026 (-$78 to $4,285) | $5,091 (-$1,101 to $12,520) | S, D, T, F, I, O, (P) |
| | | NR++ | | 9mo – TBI^G | SAT | $12,063 ($5,647 to $16,149) | $25,641 ($7,136 to $48,074) | |
| | | NR++ | SMS + reply | 9mo – TBI^C | SAT | $1,193 (-$533 to $3,037) | $4,255 (-$1,599 to $11,394) | |
| | | NR++ | | 9mo – TBI^G | SAT | $8,751 ($3,426 to $12,946) | $22,407 ($4,698 to $44,640) | |
| | | NR++ | VOT (sync.) | 9mo – TBI^C | SAT | $94,771 ($36,588 to $163,522) | $97,832 ($36,612 to $168,051) | |
| | | NR++ | | 9mo – TBI^G | SAT | $371,689 ($180,731 to $529,220) | $385,345 ($188,498 to $544,358) | |
| | | NR++ | Digital pillbox | 9mo – TBI^C | SAT | $11,392 ($3,660 to $20,894) | $14,453 ($3,545 to $26,307) | |
| | | NR++ | | 9mo – TBI^G | SAT | $48,302 ($24,062 to $68,704) | $61,956 ($27,552 to $88,431) | |
| Yang et al. (2022) | Morocco | NR++ | Digital pillbox | 6mo* | DOT+SAT | $1,834§ | | S, D, T, F, I, O |

Costs included: S = Staff, D = Drugs and Treatment, T = Travel and supplies, F = Fixed assets and technology, I = Implementation, O = Overhead, P = Patient expenses
TBI = TB infection (^C Cohort modelled based on persons who were close contacts of persons with contagious TB disease. ^G Cohort modelled as unselected persons with TBI).
++These patients were a modelled cohort. #Negative values indicate cost savings. Values in parentheses are 95% uncertainty intervals
*The model considers most patients as drug-sensitive (6 months) however patients on retreatment are modelled to be treated for 8 months, and MDR patients for 24 months
#Negative values indicate cost savings. Values in parentheses are 95% uncertainty ranges. §Calculated from available data and therefore an uncertainty range is not available.

*Table A8: Incremental cost per QALY gained with provider and societal perspectives*

| Study | Country | DAT pts. | DAT used | Intervention Duration | SoC | ICER - $/QALY gained Provider | ICER - $/QALY gained Societal | Costs included |
|---|---|---|---|---|---|---|---|---|
| Louwagie et al. (2022) | South Africa | 122 | SMS + motivational interviewing | 3mo | NR | $36,248 - QALYs gained not significant | | S, D, F, I, O |
| Saha et al. (2022) | India | 200++ | Digital pillbox | 6mo | DOT (NR) | $655& | | S, T, F, I |
| Salcedo et al. (2021) | USA | 100§ | VOT – Async. (AI) | 8mo* | DOT (Clinic & Field) | | DAT Dominant | S, D, T, F, P |

Costs included: S = Staff, D = Drugs and Treatment, T = Travel and supplies, F = Fixed assets and technology, I = Implementation, O = Overhead, P = Patient expenses
§Data inputs to model were obtained from 43 patients receiving the DAT and 73 receiving the SoC. The model then compared a cohort of 100 patients in each arm.
*The model used observed treatment completion probabilities to model successful completion or required prolongation of treatment in each arm at the end of each month (minimum 5 months, maximum 16 months). Mean treatment durations in both groups were 8 months.
&Perspective was labelled as societal in study, but all costs reported were for provider
++Saha et al. 200 patients received the intervention in a quasi-experimental study, but modeling was used for cost effectiveness based on completion probabilities found in study.

*Table A9: CHEERS 2022 Criteria checklist for included studies*

| **CHEERS 2022 Criteria** | 1 | 2 | 3 | 4 | 5 | 6 | 7 | 8 | 9 | 10 | 11 | 12 | 13 | 14 | 15 | 16 | 17 | 18 | 19 | 20 | 21 | 22 | 23 | 24 | 25 | 26 | 27 | 28 |
|---|---|---|---|---|---|---|---|---|---|---|---|---|---|---|---|---|---|---|---|---|---|---|---|---|---|---|---|---|
| Au Yeung et al. (2012) | ✓ | ✓ | ✓ | × | × | × | ✓ | ✓ | | | | | | ✓ | ✓ | ✓ | × | | | ✓ | × | ✓ | ✓ | × | × | ✓ | × | ✓ |
| Bahrainwala et al. (2020) | ✓ | ✓ | ✓ | × | ✓ | ✓ | ✓ | ✓ | ✓ | ✓ | ✓ | ✓ | ✓ | ✓ | ✓ | ✓ | ✓ | | | ✓ | × | ✓ | ✓ | ✓ | × | ✓ | ✓ | ✓ |
| Beeler Asay et al. (2020) | ✓ | ✓ | ✓ | × | × | ✓ | ✓ | ✓ | ✓ | | | | | ✓ | ✓ | ✓ | ✓ | ✓ | | ✓ | ✓ | × | ✓ | × | ✓ | ✓ | ✓ | ✓ |
| Broomhead and Mars (2012) | ✓ | ✓ | ✓ | × | ✓ | ✓ | × | ✓ | ✓ | ✓ | | | | × | ✓ | × | ✓ | | | × | × | × | ✓ | × | × | ✓ | × | ✓ |
| Buchman and Cabello (2017) | × | × | ✓ | × | × | ✓ | ✓ | × | ✓ | | | | | ✓ | × | | | | | × | × | × | × | × | × | × | ✓ | ✓ |
| Daftary et al. (2017) | | | ✓ | × | ✓ | ✓ | ✓ | × | ✓ | | | | | × | × | | | | | | × | × | ✓ | | × | ✓ | ✓ | ✓ |
| Fekadu et al. (2021) | ✓ | ✓ | ✓ | × | ✓ | ✓ | ✓ | ✓ | ✓ | ✓ | ✓ | ✓ | ✓ | ✓ | ✓ | ✓ | ✓ | | | ✓ | × | ✓ | × | ✓ | × | ✓ | ✓ | ✓ |
| Garfein et al. (2018) | × | × | ✓ | × | ✓ | × | ✓ | ✓ | ✓ | | | | | ✓ | ✓ | | ✓ | ✓ | × | × | ✓ | × | × | × | ✓ | ✓ | ✓ | ✓ |
| Gashu et al. (2021) | | | × | × | ✓ | ✓ | ✓ | × | ✓ | | | | | ✓ | × | | | | | | ✓ | | ✓ | × | ✓ | ✓ | ✓ | ✓ |
| Guo et al. (A) (2020) | | ✓ | ✓ | × | ✓ | × | × | × | | | | | | × | × | | × | | | × | × | × | ✓ | × | × | × | ✓ | ✓ |
| Guo et al. (B) (2020) | | ✓ | ✓ | × | ✓ | ✓ | ✓ | × | | | | | | ✓ | × | | ✓ | | | | ✓ | ✓ | ✓ | | ✓ | ✓ | ✓ | ✓ |
| Holzman et al (2018) | × | ✓ | ✓ | × | ✓ | ✓ | ✓ | × | × | × | | | | ✓ | × | × | ✓ | | | ✓ | ✓ | × | ✓ | ✓ | × | ✓ | ✓ | ✓ |
| Krueger et al. (2010) | × | ✓ | × | × | × | × | × | × | ✓ | × | | | | ✓ | × | ✓ | ✓ | | | × | × | × | ✓ | × | × | × | ✓ | × |
| Lam et al. (2019) | ✓ | ✓ | ✓ | × | × | ✓ | ✓ | ✓ | ✓ | | | | | ✓ | ✓ | ✓ | ✓ | | | × | × | ✓ | ✓ | × | × | ✓ | ✓ | ✓ |
| Louwagie et al. (2022) | × | × | ✓ | ✓ | ✓ | × | × | × | ✓ | | ✓ | ✓ | ✓ | ✓ | ✓ | × | × | | | × | × | × | ✓ | × | × | ✓ | ✓ | ✓ |
| Manyazewal et al. (2022) | ✓ | ✓ | ✓ | × | ✓ | × | ✓ | ✓ | ✓ | | | | | ✓ | × | | ✓ | | | ✓ | ✓ | × | × | ✓ | × | ✓ | ✓ | ✓ |
| Nsengiyumva et al. (2018) | ✓ | ✓ | ✓ | × | ✓ | ✓ | ✓ | ✓ | ✓ | ✓ | ✓ | ✓ | ✓ | ✓ | ✓ | ✓ | ✓ | ✓ | ✓ | ✓ | × | ✓ | ✓ | ✓ | × | ✓ | ✓ | ✓ |
| Nsengiyumva et al. (2023) | ✓ | ✓ | ✓ | × | × | × | × | ✓ | ✓ | ✓ | | | | ✓ | ✓ | | ✓ | | | × | ✓ | ✓ | ✓ | × | × | ✓ | ✓ | ✓ |
| Ravenscroft et al. (2020) | | ✓ | ✓ | × | ✓ | ✓ | ✓ | ✓ | | | | | | ✓ | × | ✓ | × | ✓ | ✓ | ✓ | ✓ | × | × | ✓ | ✓ | ✓ | ✓ | ✓ |
| Saha et al. (2022) | × | ✓ | × | × | ✓ | × | × | × | × | ✓ | ✓ | ✓ | ✓ | ✓ | ✓ | ✓ | ✓ | | | ✓ | × | × | ✓ | × | × | ✓ | ✓ | ✓ |
| Salcedo et al. (2021) | ✓ | ✓ | ✓ | × | ✓ | ✓ | ✓ | ✓ | ✓ | × | ✓ | ✓ | ✓ | ✓ | ✓ | ✓ | ✓ | | | ✓ | × | ✓ | ✓ | ✓ | × | ✓ | ✓ | ✓ |
| Siddiqui et al. (2019) | ✓ | ✓ | ✓ | × | ✓ | ✓ | ✓ | ✓ | ✓ | | | | | ✓ | × | ✓ | × | | | × | × | × | × | × | ✓ | ✓ | × | × |
| Story et al. (2019) | | × | | × | ✓ | × | ✓ | ✓ | ✓ | ✓ | | | | ✓ | ✓ | ✓ | | | | × | × | ✓ | ✓ | × | × | ✓ | ✓ | ✓ |
| Thompson et al. (2022) | ✓ | ✓ | ✓ | × | × | ✓ | ✓ | ✓ | ✓ | ✓ | ✓ | ✓ | × | ✓ | ✓ | ✓ | ✓ | | | ✓ | ✓ | ✓ | ✓ | ✓ | × | ✓ | ✓ | ✓ |
| Wade et al. (2012) | × | ✓ | ✓ | × | ✓ | × | ✓ | × | × | | ✓ | ✓ | ✓ | ✓ | × | ✓ | × | | | ✓ | ✓ | × | ✓ | ✓ | ✓ | ✓ | ✓ | ✓ |
| Yang et al (2022) | ✓ | ✓ | ✓ | × | ✓ | ✓ | ✓ | ✓ | ✓ | ✓ | ✓ | ✓ | ✓ | ✓ | ✓ | ✓ | ✓ | | | ✓ | × | × | ✓ | ✓ | × | ✓ | ✓ | ✓ |

Note: Only full text studies evaluated. ✓ Criterion reported. ✓ Criterion not reported. Blank cells are not applicable.

*Figure A1: Summary of incremental costs to providers. Each point represents one comparison; some studies evaluated multiple scenarios (e.g. different comparators, TB disease versus TB infection, etc.)*

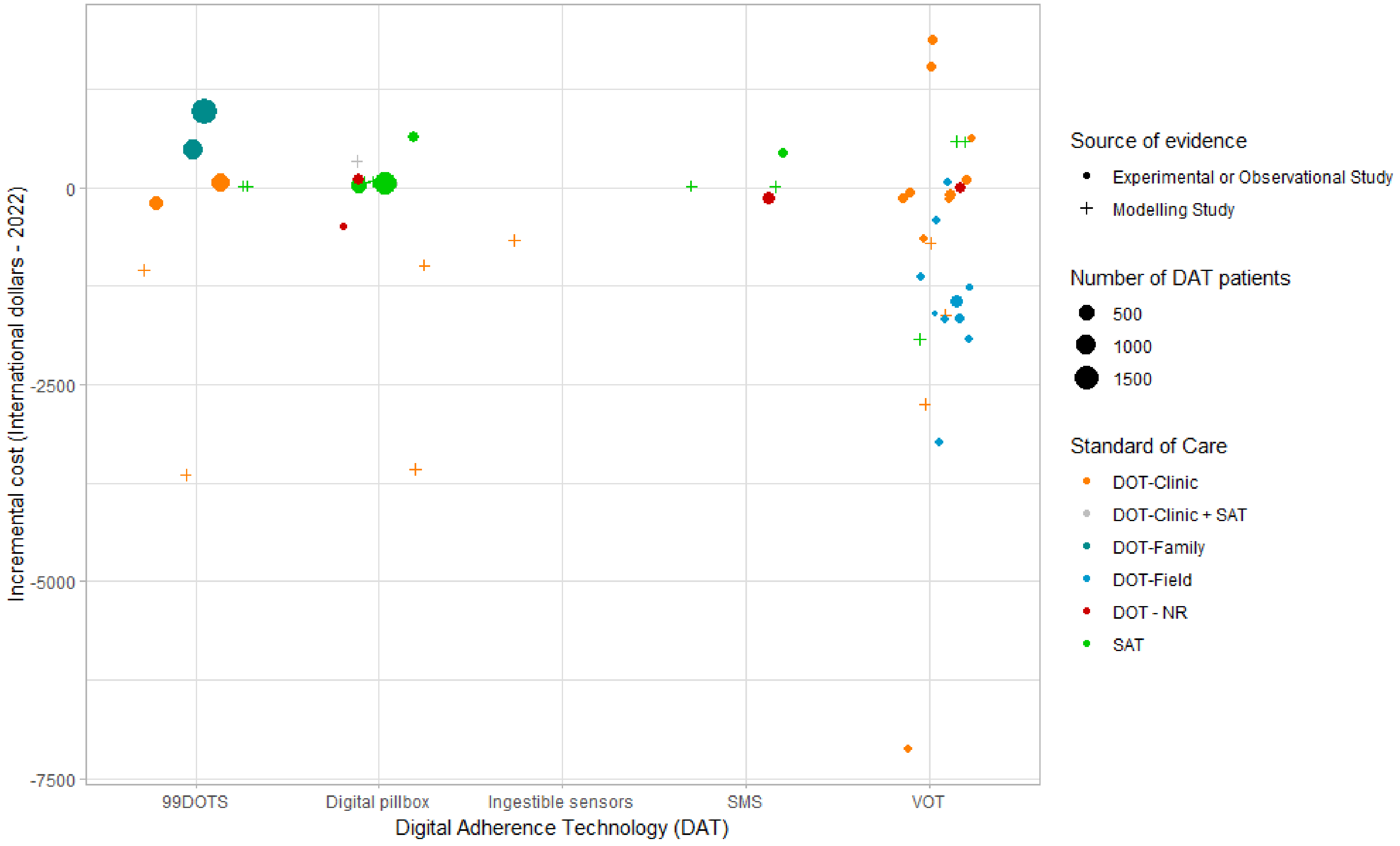


#### *S6 USD results – Sensitivity analysis*

*Table A10:* Provider costs for VOT and standard of care (SoC) – USD based on market exchange rates.

| | | | | | | Provider costs per person treated | | | |
|---|---|---|---|---|---|---|---|---|---|
| **Study** | **Country** | **DAT pts.** | **VOT Type** | **Duration** | **SOC** | **VOT** | **SoC** | **Incremental** | **Costs included** |
| Beeler Asay et al. (2020) | USA | 63 | Async. | 6mo – Calc. | DOT (Field) | $2,471 | $2,888 | -$417 | S, T, F, I |
| | | | | 6mo – Calc. | DOT (Clinic) | $2,471 | $1,856 | $616 | |
| | | 50 | Sync. | 6mo – Calc. | DOT (Field) | $1,208 | $2,888 | -$1,680 | |
| | | | | 6mo – Calc. | DOT (Clinic) | $1,208 | $1,856 | -$648 | |
| Buchman and Cabello (2017) | USA | 24 | Sync. | NR | DOT (Field) | Incremental | | -$1,256 | S, T, F |
| Fekadu et al. (2021) | USA | NR++ | Sync. | 6mo | SAT | $13,428 | $15,353 | -$1,925 | S, D, T, F |
| | | | | 6mo | DOT (Clinic) | $13,428 | $15,048 | -$1,620 | |
| Garfein et al. (2018) | USA | 225 | Async. | 6mo | DOT (Field) | $4,108 | $5,549 | -$1,441 | S, T, F |
| Guo et al. (A) (2020) | China | 199 | Sync. | NR | DOT (NR) | $5 | $11 | -$6 | S, T, F |
| Guo et al. (B) (2020) | China | 90 | Async. | 6mo | DOT (Clinic) | $8 | $44 | -$36 | T (Transport only^) |
| Holzman et al (2018) | USA | 15/vehicle± | Async. | 6mo | DOT (Field) | $775 | $2,373 | -$1,599 | S, T, F |
| Krueger et al. (2010) | USA | 57 | Sync. | 5mo | DOT (Field) | Incremental | | -$3,228 | S, T |
| Lam et al. (2019) | USA | 81 | Sync. | 6mo – Calc. | DOT (Clinic) | $1,427 | $1,319 | $108 | S, T, F |
| | | | | 6mo – Calc. | DOT (Field) | $1,427 | $3,091 | -$1,664 | |
| | | 41 | Async. | 6mo – Calc. | DOT (Clinic) | $1,168 | $1,319 | -$151 | |
| | | | | 6mo – Calc. | DOT (Field) | $1,168 | $3,091 | -$1,923 | |
| Nsengiyumva et al. (2018) | Brazil | NR++ | Sync. | 6mo – DS | DOT (Clinic) | $548 | $898 | -$351 | S, D, T, F, I, O |
| | | | | 18mo – DR | DOT (Clinic) | $10,795 | $12,155 | -$1,360 | |
| | | | | 9mo – TBI[G] | SAT | $344 | $58 | $286 | |
| | | | | 9mo – TBI[C] | SAT | $337 | $48 | $289 | |
| Nsengiyumva et al. (2023) | Moldova | 173 | Async. | 6mo – DS | DOT (Clinic) | $172 ($304) | $336 | -$164 (-$32) | S, T, F, I |
| | | 135 | Async. | 9mo – DR | DOT (Clinic) | $259 ($457) | $504 | -$245 (-$47) | |
| | Haiti | 87 | Async. | 6mo – DS | DOT (Clinic) | $1,085 ($1,154) | $414 | $671 ($740) | |
| | Philippines | 119 | Async. | 9mo - DR | DOT (Clinic) | $495 ($661) | $17 | $478 ($644) | |
| Siddiqui et al. (2019) | USA | 47 | Async. | 6mo – Calc. | DOT (Field) | $4,426 | $5,570 | -$1,145 | S, T, F |
| Story et al. (2019) | UK | 50§ | Sync. | 6mo | DOT (Clinic) | $2,400 | $8,316 | -$5,916 | S, T, F |
| Wade et al. (2012) | Australia | 47§ | Sync* | 5mo | DOT (Field) | $2,350 | $2,293 | $58 | S, T, F, O |

Costs included: S = Staff, D = Drugs and Treatment, T = Travel and supplies, F = Fixed assets and technology, I = Implementation, O = Overhead, P = Patient expenses
Calc. = Converted from per observation cost (see Methods). DS = Drug susceptible, DR = Drug resistant., Async. asynchronous, Sync. Synchronous, NR not reported. TBI = TB infection (C Cohort modelled based on persons who were close contacts of persons with contagious TB disease. G Cohort modelled as unselected persons with TBI).
Cost in parentheses are calculated with fixed costs annuitized over a 5-year useful life.
^Patients given funds to cover cost of roundtrip to clinic on public transportation §These studies observed costs from 112 patients (Story et al.) and 58 patients (Wade et al.) but modelled clinics with 50 and 47 patients respectively as base cases. These studies also provided multiple scenarios with varying patients per clinic not shown here. 5X/week DOT is shown for Story et al.
±This study observed costs from 28 patients but modelled a clinic assuming 15 patients per vehicle used in field DOT
++These patients were a modelled cohort. *Study considers 5% of patients remaining on DOT even in the DAT case.

*Table A11: Societal (or patient-only) costs for video-observed therapy (VOT) and standard of care (SoC) – USD based on market exchange rates.*

| Study | Country | DAT pts. | VOT Type | Duration | SOC | Costs per person treated | | | Costs included |
|---|---|---|---|---|---|---|---|---|---|
| | | | | | | VOT | SoC | Incremental | |
| Beeler Asay et al. (2020) | USA | 63 | Async. | 6mo | DOT (Field) | $2,707 | $7,053 | -$4,347 | S, T, F, I, P |
| | | | | 6mo | DOT (Clinic) | $2,707 | $3,274 | -$567 | |
| | | 50 | Sync. | 6mo | DOT (Field) | $1,424 | $7,053 | -$5,629 | |
| | | | | 6mo | DOT (Clinic) | $1,424 | $3,274 | -$1,849 | |
| Nsengiyumva et al. (2018) | Brazil | NR[++] | Sync. | 6mo – DS | DOT (Clinic) | $693 | $1,206 | -$513 | S, D, T, F, I, O, P |
| | | | | 18mo – DR | DOT (Clinic) | $11,250 | $13,183 | -$1,934 | |
| | | | | 9mo – TBI[G] | SAT | $461 | $161 | $299 | |
| | | | | 9mo – TBI[C] | SAT | $469 | $175 | $295 | |
| Ravenscroft et al. (2020) | Moldova | 155 | Async. | 4mo | DOT (Clinic) | $12 | $45 | -$33 | P (Transport only) |
| Salcedo et al. (2021) | USA | 100[§] | Async. (AI) | 8mo[#] | DOT (Clinic & Field) | $3,139 | $5,759 | -$2,619 | S, D, T, F, P |

Costs included: S = Staff, D = Drugs and Treatment, T = Travel and supplies, F = Fixed assets and technology, I = Implementation, O = Overhead, P = Patient expenses
AI artificial intelligence, Async. asynchronous, Sync. Synchronous, NR not reported.
TBI = TB infection (C Cohort modelled based on persons who were close contacts of persons with contagious TB disease. G Cohort modelled as unselected persons with TBI).
[++]These patients were a modelled cohort.
§Data inputs to model were obtained from 43 patients receiving the DAT and 73 receiving the SoC. The model then compared a cohort of 100 patients in each arm.
[#]The model used observed treatment completion probabilities to model successful completion or required prolongation of treatment in each arm at the end of each month (minimum 5 months, maximum 16 months). Mean treatment durations in both groups were 8 months.

*Table A12: Provider and societal costs for digital pillbox intervention and standard of care (SoC) – USD based on market exchange rates*

| | | | | | | Provider costs per person treated | | | |
|---|---|---|---|---|---|---|---|---|---|
| **Study** | **Country** | **DAT pts.** | **Co-intervention** | **Duration** | **SOC** | **DAT** | **SoC** | **Incremental** | **Costs included** |
| Bahrainwala et al. (2020) | Madagascar | 475++ | - | 6 mo | SAT | $1,404 | $1,395 | $9 | S, D, T, F, I, O |
| | | 276++ | DOT (Clinic) + SAT | 6 mo | DOT (Clinic) + SAT | $560 | $545 | $15 | |
| Broomhead & Mars (2012) | S. Africa | 24++ | DOT (NR) | 6 mo | DOT alone (NR) | $762 | $969 | -$207 | S, D, F, I |
| Mukora et al. (2022) | S. Africa | 1305^ | - | 6 mo | SAT | $50 | $29 | $22 | Abstract only |
| Nsengiyumva et al. (2018) | Brazil | NR++ | - | 6 mo – DS | DOT (Clinic) | $409 | $898 | -$490 | S, D, T, F, I, O |
| | | | | 18 mo – DR | DOT (Clinic) | $10,386 | $12,155 | -$1,768 | |
| | | | | 9 mo – TBI^G | SAT | $86 | $48 | $38 | |
| | | | | 9 mo – TBI^C | SAT | $93 | $58 | $35 | |
| Saha et al. (2022) | India | 200 | - | 6 mo | DOT (NR) | $95 | $69 | $26 | S, T, F, I |
| Yang et al. (2022) | Morocco | 206 | - | 6 mo | SAT | $396 | $155 | $241 | S, D, T, F, I, O |
| | | NR± | - | | DOT + SAT* | $740 | $615 | $125 | |
| | | | | | | **Societal costs per person treated** | | | |
| Manyazewal et al. (2022) | Ethiopia | 52 | - | 2 mo | DOT (Clinic) | $0.61 | $11 | -$10 | P |
| Nsengiyumva et al. (2018) | Brazil | NR++ | - | 6 mo – DS | DOT (Clinic) | $554 | $1,206 | -$652 | S, D, T, F, I, O, P |
| | | | | 18 mo – DR | DOT (Clinic) | $10,841 | $13,183 | -$2,342 | |
| | | | | 9 mo – TBI^G | SAT | $209 | $161 | $47 | |
| | | | | 9 mo – TBI^C | SAT | $218 | $175 | $43 | |

Costs included: S = Staff, D = Drugs and Treatment, T = Travel and supplies, F = Fixed assets and technology, I = Implementation, O = Overhead, P = Patient expenses).

DS = Drug susceptible, DR = Drug resistant, TBI = TB infection (^C Cohort modelled based on persons who were close contacts of persons with contagious TB disease. ^G Cohort modelled as unselected persons with TBI).

DOT directly observed therapy, SAT self-administered therapy, NR not reported.

++These patients were a modelled cohort. For Broomhead & Mars, while the 24 patients did receive the intervention, the costs were entirely modelled using literature values. For Bahrainwala et al. the costs are presented per diagnosed patient only (to ensure comparability between other studies)

±This study observed costs from an implementation of 206 patients but then modelled costs in a separate scenario considering costs of retreatment, MDR development & treatment, etc.

^ Target number of patients from protocol of parent RCT [1]. Actual patient numbers not reported in this abstract

*2 months of DOT (facility details not reported) followed by 4 months of SAT. The model considers most patients as drug-sensitive (6-month treatment course) however patients on retreatment are modelled to be treated for 8 months, and MDR patients for 24 months.

*Table A13: Provider and societal costs for SMS-based interventions and standard of care (SoC) – USD based on market exchange rates*

| Study | Country | DAT pts. | Co-intervention | Duration | SOC | Provider costs per person treated | | | Costs included |
|---|---|---|---|---|---|---|---|---|---|
| | | | | | | DAT | SoC | Incremental | |
| Gashu et al. (2021) | Ethiopia | 131* | DOT (Family) | 4mo | N/A | $0.16 | | | T (Airtime only) |
| Louwagie et al. (2022) | S. Africa | 122 | Motivational interviewing | 3mo | SAT | $236 | $51 | $185 | S, D, F, I, O |
| Nsengiyumva et al. (2018) | Brazil | NR++ | Patient reply | 9mo – TBI[G] | SAT | $57 | $48 | $9 | S, D, T, F, I, O |
| | | | | 9mo – TBI[C] | SAT | $63 | $58 | $5 | |
| Peng et al. (2014) | China | 234 | - | 6mo | DOT (Type not specified) | $43 | $129 | -$86 | Abstract only |
| | | | | | | **Societal costs per person treated** | | | |
| Nsengiyumva et al. (2018) | Brazil | NR++ | Patient Reply | 9mo – TBI[G] | SAT | $184 | $161 | $22 | S, D, T, F, I, O, P |
| | | | | 9mo – TBI[C] | SAT | $191 | $175 | $16 | |

Costs included: S = Staff, D = Drugs and Treatment, T = Travel and supplies, F = Fixed assets and technology, I = Implementation, O = Overhead, P = Patient expenses

Calc. = Converted from per day cost (see Methods). TBI = TB infection ([C] Cohort modelled based on persons who were close contacts of persons with contagious TB disease. [G] Cohort modelled as unselected persons with TBI).

*Target number of patients from protocol of parent RCT. Actual patient numbers not reported in this pre-print

++These patients were a modelled cohort.

*Table A14: Provider and societal costs per person for phone-based with medication sleeves intervention ("99DOTS") – USD based on market exchange rates*

| Study | Country | DAT pts. | Duration | SoC | Provider costs per person treated: DAT | SoC | Incremental | Costs included |
|---|---|---|---|---|---|---|---|---|
| Nsengiyumva et al. (2018) | Brazil | NR++ | 6mo – DS | DOT (Clinic) | $381 | $898 | -$518 | S, D, T, F, I, O |
| | | | 18mo – DR | DOT (Clinic) | $10,348 | $12,155 | -$1,806 | |
| | | | 9mo – TBI^G | SAT | $58 | $48 | $10 | |
| | | | 9mo – TBI^C | SAT | $65 | $58 | $7 | |
| Nsengiyumva et al. (2023) | Tanzania | 976 | 6mo | DOT (Family) | $163 ($174) | $0.00 | $163 ($174) | S, T, F, I |
| | Bangladesh | 719 | 6mo | DOT (Clinic) | $81 ($98) | $74 | $7 ($24) | |
| | Philippines | 396 | 6mo | DOT (Clinic) | $79 ($106) | $176 | -$97 (-$70) | |
| Thompson et al. (2022) | Uganda | 1800/year§ | 6mo# | DOT (Family) | $339 ($58) | $0 | $339 ($58) | S, T, F, I, O |
| Waswa et al (2022) | Uganda | 1086 | 6mo – Calc. | None | $27 | | | Abstract only* |
| | | | | | **Societal costs per person treated** | | | |
| Nsengiyumva et al. (2018) | Brazil | NR++ | 6mo – DS | DOT (Clinic) | $526 | $1,206 | -$680 | S, D, T, F, I, O, P |
| | | | 18mo – DR | DOT (Clinic) | $10,803 | $13,183 | -$2,380 | |
| | | | 9mo – TBI^G | SAT | $181 | $161 | $19 | |
| | | | 9mo – TBI^C | SAT | $190 | $175 | $15 | |

Costs included: S = Staff, D = Drugs and Treatment, T = Travel and supplies, F = Fixed assets and technology, I = Implementation, O = Overhead, P = Patient expenses
DS = Drug susceptible, DR = Drug resistant, TBI = TB infection (^C Cohort modelled based on persons who were close contacts of persons with contagious TB disease. ^G Cohort modelled as unselected persons with TBI). Cost in parentheses are calculated with fixed costs annuitized over a 5-year useful life.
§This study observed detailed resource unit costs from five clinics (with 81 patients) in parent RCT and then applied these costs to resource estimates from all 18 clinics in the RCT. They used these observed costs (from 891 intervention patients) and applied them to a modelled clinic with a service volume of 1800 patients per year.
#Intervention was 6 months but costs account for patients being lost to follow up. ++These patients were a modelled cohort.
*Costs described as "running costs only" (i.e. excluding start-up costs)

*Table A15: Provider and societal costs per treatment course for other DAT interventions – USD based on market exchange rates*

| Study | Country | DAT pts. | DAT used | Duration | SOC | Provider costs per person treated | | | Costs included |
|---|---|---|---|---|---|---|---|---|---|
| | | | | | | DAT | SoC | Incremental | |
| Au Yeung et al. (2012) | USA | NR[++] | Ingestible sensors | 4 mo | DOT (Clinic)* | $1,618 | $2,289 | -$671 | S, D, T, F |
| Daftary et al. (2017) | Ethiopia | 2300 (approx.) | Interactive Voice Calls | 6 mo - IPT | N/A | $49 | - | - | T |
| | | | | | | Societal costs per person treated | | | |
| Au Yeung et al. (2012) | USA | NR[++] | Ingestible sensors | 4 mo | DOT (Clinic)* | $1,680 | $3,030 | -$1,350 | S, D, T, F, P |

Costs included: S = Staff, D = Drugs and Treatment, T = Travel and supplies, F = Fixed assets and technology, I = Implementation, O = Overhead, P = Patient expenses
IPT = Isoniazid preventative therapy. *3X/week DOT is shown for Au Yeung et al. [++]These patients were a modelled cohort.

*Table A16: Incremental cost per DALY averted from provider and societal perspectives -USD based on market exchange rates*

| Study | Country | DAT pts. | DAT used | Intervention Duration | SoC | ICER - $/DALY averted # Health system | Societal | Costs included |
|---|---|---|---|---|---|---|---|---|
| Bahrainwala et al. (2020) | Madagascar | 276++ | Digital pillbox | 6mo | DOT | $36§ | | S, D, T, F, I, O |
| | | 445++ | | 6mo | SAT | $22§ | | |
| Fekadu et al. (2021) | United States | NR++ | VOT | 6mo | DOT | DAT Dominant§ | | S, D, T, F |
| | | NR++ | | 6mo | SAT | DAT Dominant& | | |
| Nsengiyumva et al. (2018) | Brazil | NR++ | 99DOTS | 9mo – TBI C | SAT | $203 ($10 to $430) | $509 (-$97 to $1,310) | S, D, T, F, I, O, (P) |
| | | NR++ | | 9mo – TBI G | SAT | $1,323 ($617 to $1786) | $2,831 ($776 to $5,320) | |
| | | NR++ | SMS + reply | 9mo – TBI C | SAT | $119 (-$55 to $300) | $425 (-$162 to $1,181) | |
| | | NR++ | | 9mo – TBI G | SAT | $966 ($370 to $1,428) | $2,473 ($517 to $4,989) | |
| | | NR++ | VOT (sync.) | 9mo – TBI C | SAT | $9,470 ($3,533 to $16,994) | $9,776 ($3,602 to $16,947) | |
| | | NR++ | | 9mo – TBI G | SAT | $41,028 ($19,832 to $58,626) | $42,536 ($20,268 to $60,316) | |
| | | NR++ | Digital pillbox | 9mo – TBI C | SAT | $1,138 ($362 to $2,097) | $1,444 ($376 to $2,705) | |
| | | NR++ | | 9mo – TBI G | SAT | $5,331 ($2,629 to $7,551) | $6,839 ($2,975 to $10,102) | |
| Yang et al. (2022) | Morocco | NR++ | Digital pillbox | 6mo* | DOT+SAT | $431&& | | S, D, T, F, I, O |

Costs included: S = Staff, D = Drugs and Treatment, T = Travel and supplies, F = Fixed assets and technology, I = Implementation, O = Overhead, P = Patient expenses
TBI = TB infection (C Cohort modelled based on persons who were close contacts of persons with contagious TB disease. G Cohort modelled as unselected persons with TBI).
++These patients were a modelled cohort. #Negative values indicate cost savings. Values in parentheses are 95% uncertainty intervals
*The model considers most patients as drug-sensitive (6 months) however patients on retreatment are modelled to be treated for 8 months, and MDR patients for 24 months
§Calculated from available data in sensitivity analyses and therefore no uncertainty range available
&95% uncertainty range via probabilistic sensitivity analysis: Reduction in DALYs (0.4385-0.0.4440) and reduction in cost ($1,924-$2,082)
&&95% uncertainty range via probabilistic sensitivity analysis: Reduction in DALYs (0.08-0.32), increased cost ($92-157)

*Table A17: Incremental cost per TB case averted with provider and societal perspectives. – USD based on market exchange rates*

| Study | Country | DAT pts. | DAT used | Intervention Duration | SoC | ICER - $/case averted[#] Provider | ICER - $/case averted[#] Societal | Costs included |
|---|---|---|---|---|---|---|---|---|
| Nsengiyumva et al. (2018) | Brazil | NR[++] | 99DOTS | 9mo – TBI[C] | SAT | $1,002 (-$39 to $2,120) | $2,519 (-$545 to $6,195) | S, D, T, F, I, O, (P) |
| | | NR[++] | | 9mo – TBI[G] | SAT | $5,969 ($2,794 to $7,990) | $12,687 ($3,531 to $23,786) | |
| | | NR[++] | SMS + reply | 9mo – TBI[C] | SAT | $590 (-$264 to $1,503) | $2,105 (-$791 to $5,637) | |
| | | NR[++] | | 9mo – TBI[G] | SAT | $4,330 ($1,695 to $6,405) | $11,086 ($2,325 to $22,087) | |
| | | NR[++] | VOT (sync.) | 9mo – TBI[C] | SAT | $46,890 ($18,103 to $80,907) | $48,405 ($18,114 to $83,147) | |
| | | NR[++] | | 9mo – TBI[G] | SAT | $183,902 ($89,421 to $261,845) | $190,659 ($93,264 to $269,334) | |
| | | NR[++] | Digital pillbox | 9mo – TBI[C] | SAT | $5,636 ($1,811 to $10,338) | $7,151 ($1,754 to $13,016) | |
| | | NR[++] | | 9mo – TBI[G] | SAT | $23,899 ($11,905 to $33,993) | $30,654 ($13,632 to $43,753) | |
| Yang et al. (2022) | Morocco | NR[++] | Digital pillbox | 6mo* | DOT+SAT | $690[§] | | S, D, T, F, I, O |

Costs included: S = Staff, D = Drugs and Treatment, T = Travel and supplies, F = Fixed assets and technology, I = Implementation, O = Overhead, P = Patient expenses
TBI = TB infection ([C] Cohort modelled based on persons who were close contacts of persons with contagious TB disease. [G] Cohort modelled as unselected persons with TBI).
[++]These patients were a modelled cohort. [#]Negative values indicate cost savings. Values in parentheses are 95% uncertainty intervals
*The model considers most patients as drug-sensitive (6 months) however patients on retreatment are modelled to be treated for 8 months, and MDR patients for 24 months
[#]Negative values indicate cost savings. Values in parentheses are 95% uncertainty ranges. [§]Calculated from available data and therefore an uncertainty range is not available.

*Table A18: Incremental cost per QALY gained with provider and societal perspectives – USD based on market exchange rates*

| Study | Country | DAT pts. | DAT used | Intervention Duration | SoC | ICER - $/QALY gained Provider | ICER - $/QALY gained Societal | Costs included |
|---|---|---|---|---|---|---|---|---|
| Louwagie et al. (2022) | South Africa | 122 | SMS + motivational interviewing | 3mo | NR | $15,458- QALYs gained not significant | | S, D, F, I, O |
| Saha et al. (2022) | India | 200[++] | Digital pillbox | 6mo | DOT (NR) | $168[&] | | S, T, F, I |
| Salcedo et al. (2021) | USA | 100[§] | VOT – Async. (AI) | 8mo* | DOT (Clinic & Field) | | DAT Dominant | S, D, T, F, P |

Costs included: S = Staff, D = Drugs and Treatment, T = Travel and supplies, F = Fixed assets and technology, I = Implementation, O = Overhead, P = Patient expenses
§Data inputs to model were obtained from 43 patients receiving the DAT and 73 receiving the SoC. The model then compared a cohort of 100 patients in each arm.
*The model used observed treatment completion probabilities to model successful completion or required prolongation of treatment in each arm at the end of each month (minimum 5 months, maximum 16 months). Mean treatment durations in both groups were 8 months.
[&]Perspective was labelled as societal in study, but all costs reported were for provider
[++]Saha et al. 200 patients received the intervention in a quasi-experimental study, but modeling was used for cost effectiveness based on completion probabilities found in study.

One study compared synchronous VOT to field DOT and estimated it would cost $1.17 per additional treatment observation accomplished (95% confidence interval: $0.45-$2.00) to implement VOT in a clinic of 47 patients [2].

Another study compared an intervention involving medication sleeves and phone registration of doses (99DOTS) to family DOT in Uganda, and estimated it would cost $397 per additional treatment success (95% confidence interval: $245-$697) to implement the intervention in a clinic over 5 years [3].

*Table A19: Provider costs for VOT and standard of care (SoC) for studies with multiple sites - USD*

| Study | Country | Site | DAT pts. | VOT Type | Duration | SOC | Provider costs per person treated: VOT | SoC | Incremental | Costs included |
|---|---|---|---|---|---|---|---|---|---|---|
| Beeler Asay et al. (2020) | USA | NYC | 186/year | Async. | 6mo – Calc. | DOT (Field) | $1,394 | $3,375 | -$1,980 | S, T, F, I |
| | | NYC | 186/year | Async. | 6mo – Calc. | DOT (Clinic) | $1,394 | $1,054 | $340 | |
| | | NYC | 399/year | Sync. | 6mo – Calc. | DOT (Field) | $1,208 | $3,375 | -$2,167 | |
| | | NYC | 399/year | Sync. | 6mo – Calc. | DOT (Clinic) | $1,208 | $1,054 | $154 | |
| | | RI | 9/year | Async. | 6mo – Calc. | DOT (Field) | $4,624 | $3,139 | $1,485 | |
| | | SF | 62/year | Async. | 6mo – Calc. | DOT (Field) | $1,491 | $2,449 | -$959 | |
| | | SF | 62/year | Async. | 6mo – Calc. | DOT (Clinic) | $1,491 | $3,942 | -$2,452 | |
| Krueger et al. (2010) | USA | Pierce | 41 | Sync.* | 5mo | DOT (Field) | Incremental | | -$3,171 | S, T |
| | | Squamish | 16 | Sync.* | 5mo | DOT (Field) | Incremental | | -$3,373 | |

Note: Beeler Asay et al. did not provide a specific site breakdown on the number of patients that made up the combined sample of 63 patients for asynchronous and 57 patients for synchronous VOT. They did provide however annual patient volumes for each site.
*Study considers savings in patients that started on DOT and were switched to VOT for variable portions of their treatment (average of 20 weeks on VOT for all patients)

Beeler Asay et al. had 3 study sites in the United States described as New York City, Rhode Island and San Francisco. The Rhode Island site had a much smaller case load than the other sites and so fixed software costs for asynchronous VOT were proportionally much higher and represented over 80% of the per patient cost. Societal costs were not broken down by site for this study. While costs were presented in the study on a per session basis, the costs were converted to per patient using the process described in *Methods*.

*Table A20: Provider costs for VOT and DOT on a per observation basis – USD based on market exchange rates*

| Study | Country | DAT pts. | VOT Type | SOC | Provider costs per observation | | | Costs included |
|---|---|---|---|---|---|---|---|---|
| | | | | | **VOT** | **SoC** | **Incremental** | |
| Beeler Asay et al. (2020) | USA | 63 | Async. | DOT (Field) | $13.58 | $22.22 | -$8.64 | S, T, F, I |
| | | | | DOT (Clinic) | $13.58 | $14.27 | -$0.69 | |
| | | 50 | Sync. | DOT (Field) | $6.64 | $22.22 | -$15.58 | |
| | | | | DOT (Clinic) | $6.64 | $14.27 | -$7.64 | |
| Fekadu et al. (2021) | USA | NR++ | Sync. | DOT (Clinic) | $79.93 | $125.40 | -$45.47 | S, D, T, F |
| Garfein et al. (2018) | USA | 225 | Async. | DOT (Field) | $22.57 | $42.68 | -$20.11 | S, T, F |
| Guo et al. (B) (2020) | China | 90 | Async. | DOT (Clinic) | $0.09 | $0.64 | -$0.55 | T (Transport only^) |
| Holzman et al (2018) | USA | 15/vehicle± | Async. | DOT (Field) | $5.96 | $18.26 | -$12.30 | S, T, F |
| Krueger et al. (2010) | USA | 57 | Sync. | DOT (Field) | Incremental | | -$41.62 | S, T |
| Lam et al. (2019) | USA | 81 | Sync. | DOT (Clinic) | $7.84 | $10.14 | -$2.30 | S, T, F |
| | | | | DOT (Field) | $7.84 | $23.78 | -$15.94 | |
| | | 41 | Async. | DOT (Clinic) | $6.41 | $10.14 | -$3.73 | |
| | | | | DOT (Field) | $6.41 | $23.78 | -$17.36 | |
| Nsengiyumva et al. (2018) | Brazil | NR++ – DS | Sync. | DOT (Clinic) | $7.02 | $11.52 | -$4.49 | S, D, T, F, I, O |
| | | NR++ – DR | Sync. | DOT (Clinic) | $46.13 | $51.94 | -$5.81 | |
| Nsengiyumva et al. (2023) | Moldova | 173 – DS | Async. | DOT (Clinic) | $1.69 ($0.96) | $2.80 | -$1.11 (-$1.84) | S, T, F, I |
| | | 135 – DR | Async. | DOT (Clinic) | $1.69 ($0.96) | $2.80 | -$1.11 (-$1.84) | |
| | Haiti | 87 | Async. | DOT (Clinic) | $6.41 ($6.03) | $3.45 | $2.96 ($2.58) | |
| | Philippines | 119 | Async. | DOT (Clinic) | $2.45 ($1.83) | $0.09 | $2.35 ($1.74) | |
| Siddiqui et al. (2019) | USA | 47 | Async. | DOT (Field) | $24.32 | $42.85 | -$18.53 | S, T, F |
| Story et al. (2019) | UK | 50§ | Sync. | DOT (Clinic) | $13.19 | $63.97 | -$50.78 | S, T, F |
| Wade et al. (2012) | Australia | 47§ | Sync** | DOT (Field) | $16.67 | $24.92 | -$8.25 | S, T, F, O |

Costs included: S = Staff, D = Drugs and Treatment, T = Travel and supplies, F = Fixed assets and technology, I = Implementation, O = Overhead, P = Patient expenses
DS = Drug susceptible, DR = Drug resistant. Cost in parentheses are calculated with fixed costs annuitized over a 5-year useful life.
§These studies observed costs from 112 patients (Story et al.) and 58 patients (Wade et al.) but modelled clinics with 50 and 47 patients respectively as base cases. These studies also provided multiple scenarios with varying patients per clinic not shown here. 5X/week DOT is shown for Story et al.
^Patients given funds to cover cost of roundtrip to clinic on public transportation
±This study observed costs from 28 patients but modelled a clinic assuming 15 patients per vehicle used in field DOT. ++These patients were a modelled cohort.
**Study considers 5% of patients remaining on DOT even in the DAT case

*Table A21:* Provider costs for synchronous and asynchronous VOT on a per observation basis – USD based on market exchange rates

| Study | Country | Async. pts. | Sync. Pts. | Setting | Provider costs per observation | | | Costs Included |
|---|---|---|---|---|---|---|---|---|
| | | | | | Asynchronous | Synchronous | Incremental | |
| Lam et al. (2019) | USA | 81 | 41 | **New York City** | **$6.41** | **$7.84** | **-$1.43** | S, T, F |
| Beeler Asay et al. (2020) | USA | 257/year | 399/year | All sites | $13.58 | $6.64 | $6.94 | S, T, F, I |
| | | 186/year | 399/year | **New York City** | **$7.66** | **$6.64** | **$1.02** | |
| | | 62/year | - | San Francisco | $8.19 | - | | |
| | | 9/year | - | Rhode Island | $25.40 | - | | |

Costs included: S = Staff, D = Drugs and Treatment, T = Travel and supplies, F = Fixed assets and technology, I = Implementation, O = Overhead, P = Patient expenses
Note: Beeler Asay et al. did not report a site breakdown on the number of patients that made up the combined sample of 63 patients for asynchronous and 57 patients for synchronous VOT. They provided annual patient volumes for each site

*Figure A2:: Summary of incremental costs to providers – USD based on market exchange rates*

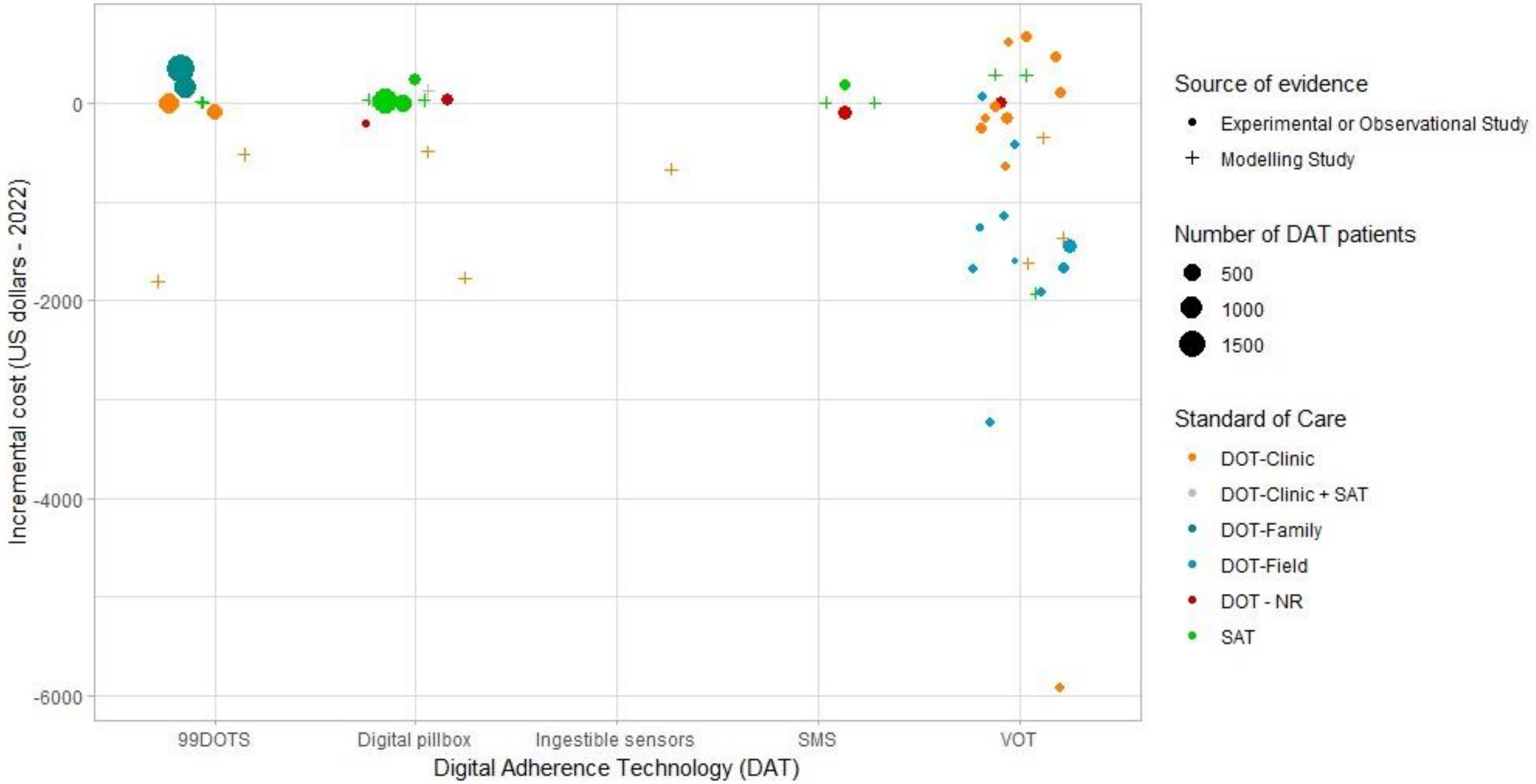

*Table A22:* Cost multipliers used to express estimates in common units

| **Authors** | **Year** | **Country** | **Currency & Year Reported** | **Local currency** | **A. Exchange rate multiplier for local currency** | **B. Local Inflation[1]** | **C. Conversion to 2022-$I[2]** | **D. Conversion to $USD[3]** |
|---|---|---|---|---|---|---|---|---|
| Au Yeung et al. [4] | 2012 | United States | USD-2011 | USD | 1.00 | 1.29 | 1.00 | 1.00 |
| Bahrainwala et al. [5] | 2020 | Madagascar | USD-2019 | MGA | 3618.32^ | 1.21 | 1199.19 | 4096.12 |
| Beeler Asay et al. [6] | 2020 | United States | USD-2017 | USD | 1.00 | 1.18 | 1.00 | 1.00 |
| Broomhead & Mars [7] | 2012 | South Africa | USD-2005 | ZAR | 6.60 | 2.68 | 6.98 | 16.36 |
| Buchman & Cabello [8] | 2017 | United States | USD-2017* | USD | 1.00 | 1.18 | 1.00 | 1.00 |
| Daftary et al. [9] | 2017 | Ethiopia | USD-2017* | ETB | 23.87^ | 2.35 | 17.03 | 51.76 |
| Fekadu et al. [10] | 2021 | United States | USD-2021 | USD | 1.00 | 1.07 | 1.00 | 1.00 |
| Garfein et al. [11] | 2018 | United States | USD-2017 | USD | 1.00 | 1.18 | 1.00 | 1.00 |
| Gashu et al. [12] (pre-print) | 2021 | Ethiopia | USD-2021* | ETB | 15.00 | 1.31 | 17.03 | 51.76 |
| Guo et al. (A) [13] | 2020 | China | CNY-2020* | CNY | 1.00 | 1.08 | 4.10 | 6.74 |
| Guo et al. (B) [14] | 2020 | China | CNY-2020* | CNY | 1.00 | 1.08 | 4.10 | 6.74 |
| Holzman et al. [15] | 2018 | United States | USD-2018* | USD | 1.00 | 1.15 | 1.00 | 1.00 |
| Krueger et al. [16] | 2010 | United States | USD-2010* | USD | 1.00 | 1.32 | 1.00 | 1.00 |
| Lam et al. [17] | 2019 | United States | USD-2016 | USD | 1.00 | 1.20 | 1.00 | 1.00 |
| Louwagie et al. [18] | 2022 | South Africa | USD-2019 | ZAR | 14.45 | 1.18 | 6.98 | 16.36 |
| Manyazewal et al. [19] | 2022 | Ethiopia | ETB-2021 | ETB | 1.00 | 1.31 | 17.03 | 51.76 |
| Mukora et al. [20] (abstract) | 2022 | South Africa | USD-2022* | ZAR | 16.45 | 1.00 | 6.98 | 16.36 |
| Nsengiyumva et al. [21] | 2018 | Brazil | USD-2016 | BRL | 3.49^ | 1.43 | 2.56 | 5.16 |
| Nsengiyumva et al. [22] (pre-print) | 2023 | Bangladesh | USD-2022 | BDT | 91.75^ | 1.00 | 32.11 | 91.75 |
| | | Haiti | USD-2022 | HTG | 115.63^ | 1.00 | 55.74 | 115.63 |
| | | Moldova | USD-2022 | MDL | 18.90^ | 1.00 | 6.64 | 18.90 |
| | | Philippines | USD-2022 | PHP | 54.48^ | 1.00 | 18.78 | 54.48 |
| | | Tanzania | USD-2022 | TZS | 2300.58^ | 1.00 | 853.89 | 2300.58[&] |
| Peng et al. [23] (abstract) | 2014 | China | USD-2014* | CNY | 6.14^ | 1.21 | 4.10 | 6.74 |
| Ravenscroft et al. [24] | 2020 | Moldova | MDL-2020* | MDL | 1.00 | 1.22 | 6.64 | 18.90 |
| Saha et al. [25] | 2022 | India | USD-2020 | INR | 79.58 | 1.13 | 20.14 | 78.60 |
| Salcedo et al. [26] | 2021 | United States | USD-2017 | USD | 1.00 | 1.18 | 1.00 | 1.00 |
| Siddiqui et al. [27] | 2019 | United States | USD-2014[&] | USD | 1.00 | 1.22 | 1.00 | 1.00 |
| Story et al. [28] | 2019 | UK | GBP-2016 | GBP | 1.00 | 1.18 | 0.67 | 0.81 |
| Thompson et al. [3] | 2022 | Uganda | USD-2019 | UGX | 3704.00 | 1.12 | 1298.23 | 3689.82 |
| Wade et al. [2] | 2012 | Australia | AUD-2012* | AUD | 1.00 | 1.28 | 1.51 | 1.44 |
| Waswa et al. [29] (abstract) | 2022 | Uganda | USD-2021 | UGX | 3587.05^ | 1.07 | 1298.23 | 3689.82 |
| Yang et al. [30] | 2022 | Morocco | USD-2018 | MAD | 9.12 | 1.11 | 3.82 | 10.16 |

Notes: Total adjustment factor (PPP - $I) = A * B * C. Total adjustment factor ($USD) = A * B * D
1. Inflation in local currency using GDP implicit deflator from IMF [31] from the currency year to 2022
2. Conversion using PPP from IMF dataset [31]
3. Using average exchange rates for 2022 as reported by the IMF [32]. [&]Average of first 8 months of 2022 since entire period was unavailable for Tanzania
*Currency year assumed to be publication year, ^Exchange rate to local currency not explicitly reported. Average market exchange rates for the currency year used (obtained from IMF dataset [32]) [&]Not explicitly reported but referenced cost inputs from 2014.

#### *S7 References*

12. Gashu KD, Gelaye KA, Tilahun B. Feasibility, acceptability and challenges of phone reminder system implementation for tuberculosis pill refilling and medication in

Northwest Ethiopia. 2021;

13. Guo P, Qiao W, Sun Y, Liu F, Wang C. Telemedicine technologies and tuberculosis management: a randomized controlled trial. Telemed e-Health. 2020;26(9):1150–6.

14. Guo X, Yang Y, Takiff HE, Zhu M, Ma J, Zhong T, et al. A comprehensive app that improves tuberculosis treatment management through video-observed therapy: usability study. JMIR mHealth uHealth. 2020;8(7):e17658.

15. Holzman SB, Zenilman A, Shah M. Advancing patient-centered care in tuberculosis management: a mixed-methods appraisal of video directly observed therapy. In: Open forum infectious diseases. Oxford University Press US; 2018. p. ofy046.

23. Peng H, Lu W, Xu W. Mobile phone text messaging for promoting adherence to anti-tuberculosis treatment: a community-randomised trial in Jiangsu, China. In: 45th World

Conference on Lung Health of the International Union Against Tuberculosis and Lung Disease (The Union). International Union Against Tuberculosis and Lung Disease (The Union); 2014. p. S213-214.
